# Distinct characteristics of lymphoid and myeloid clonal hematopoiesis in World Trade Center first responders

**DOI:** 10.1101/2024.08.01.24311359

**Authors:** Myvizhi Esai Selvan, Pei-Fen Kuan, Xiaohua Yang, John Mascarenhas, Robert J. Klein, Benjamin J. Luft, Paolo Boffetta, Zeynep H. Gümüş

## Abstract

Clonal hematopoiesis of indeterminate potential (CHIP) represents the presence of clonal somatic mutations in blood cells in otherwise healthy individuals. While CHIP is known to increase risk for hematologic malignancies and cardiovascular disease, its association with airborne carcinogens remains largely unknown. We investigated CHIP mutations in 9/11 World Trade Center (WTC) disaster responders (n=350), who experienced substantial exposure to a complex mix of airborne carcinogens. Ultra-deep whole-exome sequencing at 250X was performed on banked blood samples. We characterized CHIP mutations and their associations with clinical factors (age, ancestry, gender, body mass index, cardiovascular disease, stroke), laboratory parameters (peripheral blood counts), mental and cognitive assessments, exposure data, and HLA zygosity. Statistical methods included Fisher’s exact test, Wilcoxon rank sum test, and multivariate logistic regression. Findings were compared to unexposed controls (n=293) analyzed using identical methods. Among WTC participants, CHIP prevalence was 34.2%, with myeloid-CHIP (M-CHIP) at 16.2% and lymphoid-CHIP (L-CHIP) at 21.4%. M-CHIP prevalence correlated positively with age (*p*=0.02), smoking history (*p=*0.01), and lower platelet counts (*p*=0.03). The most frequent M-CHIP mutations were in *DNMT3A, TET2, PPM1D*, while L-CHIP mutations were in *EEF1A1, DDX11* and *KMT2D*. Notably, *DDX11* mutations were associated with lower Montreal Cognitive Assessment scores (*p*=6.57e-03). Comparison with unexposed controls demonstrated higher CHIP prevalence in WTC responders, particularly in those 55 or younger. Study highlights the potential utility of deep sequencing for CHIP detection with clinical, laboratory and exposure data to develop personalized risk-adapted screening programs for cancer and other CHIP-related conditions in individuals exposed to airborne carcinogens.

---

More than two decades after the 9/11 attacks, long-term health consequences for World Trade Center (WTC) rescue and recovery workers continue to emerge. Over 91,000 individuals participated in rescue, recovery, debris cleanup, and restoration of essential services. These first responders, including firefighters, police officers, paramedics, engineers, steel workers, railway tunnel workers, telecommunications specialists, sanitation workers, medical examiners, and volunteers, had no prior training in civil disaster response. These responders faced unprecedented exposure to a complex mix of known or suspected airborne carcinogens, raising concerns regarding their long-term cancer risk. WTC responders inhaled a toxic blend including benzene, formaldehyde, asbestos, silica, cement dust, glass fibers, heavy metals, polycyclic aromatic hydrocarbons, polychlorinated biphenyls, polychlorinated dibenzofurans, and dioxins^1^. While the carcinogenicity of these individual compounds is well-documented, their collective impact on hematologic malignancy risk remains poorly characterized, creating a critical knowledge gap.

A key mechanism potentially linking environmental exposures to cancer development is clonal hematopoiesis^2^, where a subpopulation of blood cells harbors clonal genomic mutations associated with blood cancers, without detectable hematologic disorders or unexplained persistent cytopenia. Clonal hematopoiesis of indeterminate potential (CHIP) involves clonal subpopulations carrying a point mutation or short insertion/deletion with a variant allele fraction (VAF) of at least 2% in genes recurrently mutated in hematologic malignancies. While most research has focused on myeloid lineage CHIP (M-CHIP), lymphoid lineage CHIP (L-CHIP) also occurs and may contribute to lymphoid malignancy risk, though it remains understudied. Beyond cancer risk, CHIP is associated with cytopenias, cardiovascular disease, infection susceptibility and all-cause mortality, making it a potentially important biomarker for multiple health conditions relevant to WTC responders.

To characterize the full spectrum of CHIP mutations in WTC responders, we performed ultra-deep Whole Exome Sequencing (WES) at 250X on 350 blood samples from 345 participants from the World Trade Center Health Program (WTCHP) General Responders Cohort (GRC) cohort aged 48-90 years (median, 59 years), without a history of hematologic malignancy at enrollment (Figure 1A, Table 1). After rigorous quality control, we identified M-CHIP and L-CHIP mutations (Figure 1B, Supplementary Table S1) and analyzed their associations with age, ancestry, WTC exposure, HLA zygosity, and various clinical, laboratory, mental and cognitive metrics. CHIP prevalence was also compared to an unexposed New York City control cohort (n=293; Supplementary Table S2). Detailed methodology is provided in the supplemental materials.

**Table 1.** Characteristics of the World Trade Center Health Program (WTCHP) General Responders Cohort (GRC) members selected for WES in this study (the WTC cohort), which, after sample QC that removed duplicates, totaled 345 individuals.

| Phenotype | Category | # Samples (%) |
| --- | --- | --- |
| Sex (M/F =12.3) | Male (M) | 319 (92.5%) |
|  | Female (F) | 26 (7.5%) |
| Smoking status | Current smoker | 15 (4.3%) |
|  | Previous smoker | 154 (44.6%) |
|  | Never smoker | 176 (51.0%) |
| Race | White/ Caucasian | 314 (91.0%) |
|  | Black/ African-American | 12 (3.5%) |
|  | Other race | 13 (3.8%) |
|  | Unknown | 6 (1.7%) |
| Age (yrs)<br>Median= 59<br>SD= 5.90 | ≤ 55 | 27 (7.8%) |
|  | 56 - 60 | 171 (49.6%) |
|  | 61 - 65 | 84 (24.3%) |
|  | 66 - 70 | 32 (9.3%) |
|  | > 70 | 31 (9.0%) |
| Derived exposure level | Very high | 15 (4.3%) |
|  | High | 55 (15.9%) |
|  | Intermediate | 210 (60.9%) |
|  | Low | 50 (14.5%) |
|  | Very low | 6 (1.7%) |
|  | Missing/ No record | 9 (2.6%) |

**Figure 1.**
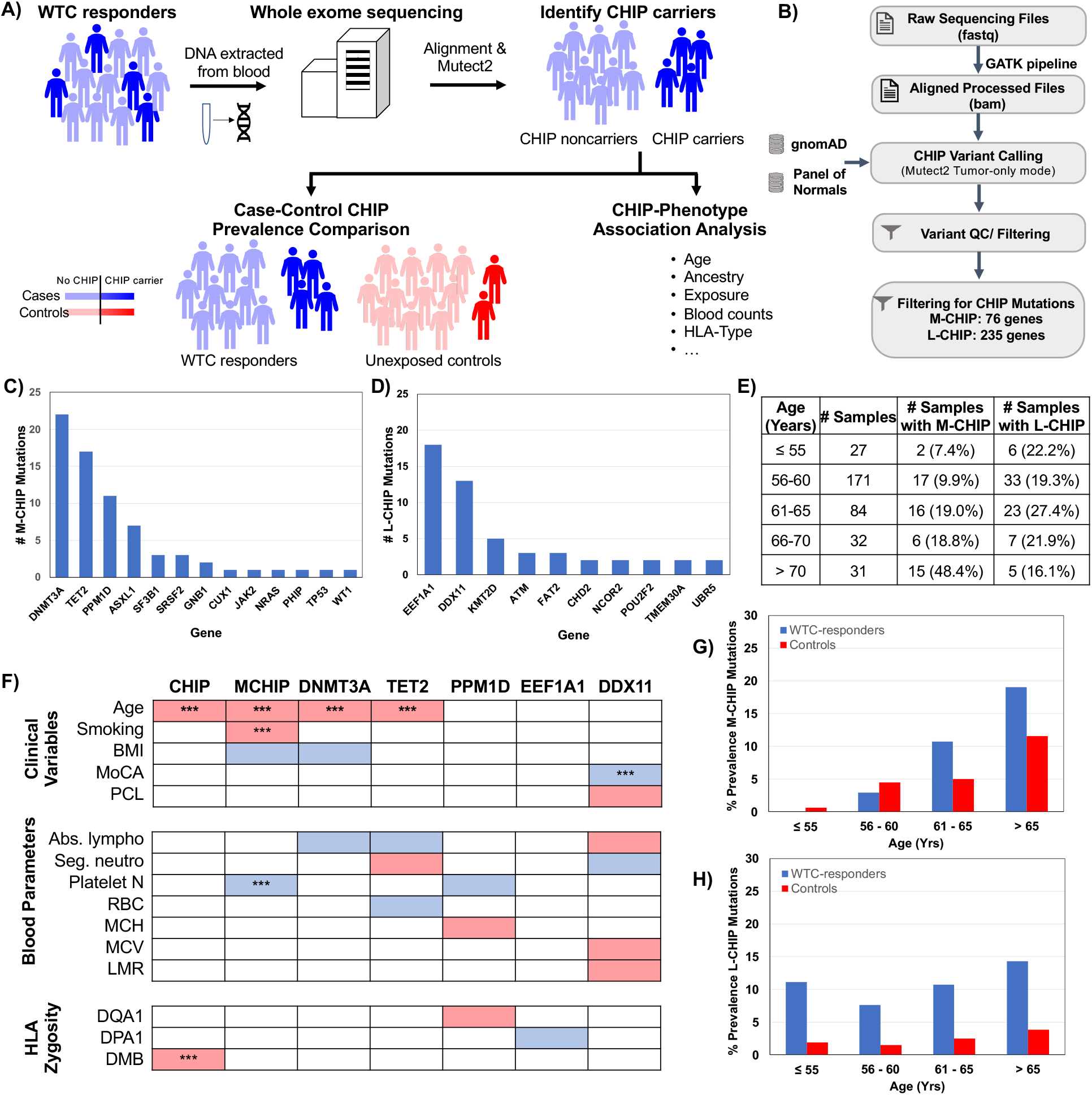
**A)** Study outline - To understand CHIP prevalence in WTC debris-exposed first responders, we collected blood from participants, extracted DNA, and performed deep WES at 250X on 350 samples. Next, we aligned the exomes to the human genome and called somatic variants using Mutect2 to identify CHIP carriers, the specific CHIP mutations and VAF. Additionally, we compared CHIP prevalence in WTC debris-exposed first responders to healthy controls from the New York City area. Finally, we associated CHIP prevalence with respect to age, ancestry, exposure, cognitive/metal data, HLA zygosity, blood counts and other clinical variables; **B)** WES data analysis pipeline to identify CHIP mutations; **C)** Number of M-CHIP mutations observed in each gene; **D)** Number of L-CHIP mutations observed in top genes (#Mutations > 1); **E)** Prevalence of M-CHIP and L-CHIP mutations; **F)** CHIP-phenotype associations - Heatmap shows significant associations between overall CHIP, M-CHIP, M-CHIP driven by *DNMT3A, TET2, PPM1D* mutations and L-CHIP driven by *EEF1A1, DDX11* mutations versus clinical variables, blood parameters and HLA zygosity. Blue and red correspond to significant (*p* ≤ 0.05) negative (OR < 1) and positive (OR > 1) correlation. White represents absence of significant correlation. Asterisk represents significant associations (*p* ≤ 0.05) in the multivariate regression analysis; **G)** M-CHIP mutation prevalence in 345 WTC debris-exposed individuals (blue) and 293 unexposed controls (red) as a function of age; **H)** L-CHIP mutation prevalence in 345 WTC debris-exposed individuals (blue) and 293 unexposed controls (red) as a function of age. A statistically significant difference in prevalence was observed between the two groups (*p* = 0.031).

We found that 34.2% (118/345) of WTC participants harbored at least one CHIP mutation. 16.2% harbored M-CHIP mutations (56 participants, 71 mutations, 13 genes), and 21.4% carried L-CHIP mutations (74 participants, 85 mutations, 43 genes) (Supplementary Table S3). Clonal complexity (≥2 mutations) was present in 7.0% (24/345), and 3.5% (12 participants) carried both M-CHIP and L-CHIP mutations. The majority (>80%) of CHIP-positive participants harbored only a single mutation (Supplementary Figure S1). Among M-CHIP mutations, 39% were non-synonymous, 30% were stop-gain, 23% were frameshift deletions, and the remainder were frameshift insertions and splicing variants. The majority of L-CHIP mutations (87%) were non-synonymous, 9% were stop-gain, and the rest were frameshift insertions/deletions (Supplementary Figure S1). The most frequently mutated M-CHIP genes were *DNMT3A, TET2, PPM1D*, and *ASXL1*, and L-CHIP genes were *EEF1A1, DDX11, KMT2D, ATM*, and *FAT2* (Figure 1C,D). Mutations in *EEF1A1* (21%) and *DDX11* (15%), particularly at (*EEF1A1*:E293K, *EE1F1A*:V315L and *DDX11*:P368S) drove the L-CHIP signal. Among all CHIP mutations, *TET2* exhibited the highest VAF (Supplementary Figure S2).

Consistent with prior studies^3^, M-CHIP prevalence increased with age, especially for *DNMT3A* and *TET2*, which remained significant in multivariate logistic regression (Figure 1E-F, Supplementary Table S4). Median ages for CHIP-positive, M-CHIP, *DNMT3A*-mutant, and *TET2*-mutant participants were 61, 62, 66.5 and 64 years respectively, compared to 59 years for CHIP-negative participants. Smoking history positively associated with M-CHIP in both univariate and multivariate regression analyses. Associations between BMI and M-CHIP, particularly BMI and *DNMT3A* may require larger sample sizes to clarify.

As WTC responders age, they face increased risks for neurocognitive and motor dysfunction that resembles neurodegenerative diseases, along with cortical atrophy and cognitive impairment at midlife, associated with both physical exposures at the WTC site, and chronic PTSD. Notably, participants with *DDX11* mutations showed significantly higher PTSD Checklist (PCL) scores (indicating greater PTSD severity) and lower Montreal Cognitive Assessment (MoCA) scores (indicating mild cognitive impairment) (Figure 1F). MoCA score association remained significant in multivariate logistic regression. The median MoCA score for participants with *DDX11* mutations was 22, falling within the range indicative of mild cognitive impairment. Given that *DDX11* is a helicase involved in altering RNA secondary structure, and helicase dysfunction has been implicated in neurodegeneration, these findings suggest a link between CHIP and neurocognitive outcomes worthy of further investigation.

In laboratory parameters (Figure 1F, Supplementary Figure S3), M-CHIP positive cases showed lower platelet counts in both univariate and multivariate regression models compared to CHIP-negative cases. *DNMT3A* and *TET2* mutation carriers had lower absolute lymphocyte counts, and *TET2* mutation also associated with lower red blood cell (RBC) counts and higher segmented neutrophils. *PPM1D* mutations correlated with lower platelet counts and higher mean corpuscular hemoglobin (MCH). *DDX11* mutation carriers demonstrated elevated absolute lymphocytes counts, mean corpuscular volume (MCV), lymphocyte-to-monocyte ratio (LMR), and lower segmented neutrophil counts.

Comparative analyses against UK Biobank data^4^ highlighted both convergent (e.g. *TET2* with lower lymphocyte counts and *PPM1D* with lower platelet counts) and divergent (e.g. M-CHIP associations with lower platelet counts and *TET2* associations with elevated segmented neutrophil counts) patterns. However, multivariate analysis adjusting for age and other factors attenuated most associations. Differences may reflect WTC-debris exposure or methodological factors.

Recent population-level studies suggest that abnormal complete blood counts (CBC) may predict future CHIP development, and CHIP-positive individuals with abnormal myeloid or lymphoid counts face highest risks for corresponding malignancies. Thus, CHIP-positive WTC responders with elevated myeloid and/or lymphoid blood counts may represent a high-priority subgroup for enhanced surveillance. Given the emerging interest in targeting inflammatory pathways using inhibitors of IL-1β, *NLRP3*, and *IRAK1* in CHIP, such interventions in high-risk CHIP-positive WTC participants may help prevent or delay overt myeloid neoplasms, CVD, major bleeding, and infection.

Next, cognizant of CHIP association with immune dysfunction, we examined Human Leukocyte Antigen (HLA) type (both classes I and II) zygosity in relation to CHIP. The heterozygote advantage hypothesis suggests that heterozygous HLA genotypes may confer broader immune response capabilities than homozygous HLA genotypes. CHIP prevalence was statistically significantly associated with *HLA-DMB* homozygosity, which remained significant in multivariate logistic regression, with weaker associations between *PPM1D* and *HLA-DQA1* homozygosity, and inverse associations between *EEF1A1* and *HLA-DPA1* homozygosity. These findings implicate HLA Class II antigen presentation in CHIP risk, supporting a role for immune-genetic interactions.

Note that cross-study comparisons of CHIP prevalence remain challenging due to evolving definitions and lack of standardized analytical pipelines. The 2022 World Health Organization CHIP definition includes somatic mutations in myeloid malignancy-associated genes at VAF ≥ 2% (≥4% for X-linked gene mutations in males) in individuals without hematologic disorders or unexplained cytopenia. However, no guidelines exist on corresponding read depth requirements, as mutations with a true VAF of 2% are more likely missed at 30X than 250X depth. Studies also vary on CHIP variants/genes, without international consensus, and no established ‘gold standard’ analysis pipelines exist for CHIP detection. Technical artifact filtering also varies between studies, further complicating cross-study comparisons. Therefore, we compared CHIP prevalence to healthy, unexposed controls from the New York area using the same CHIP calling pipeline. We observed a generally higher CHIP prevalence in the WTC cohort (Figure 1G,H, Supplementary Table S5). After down-sampling for comparable sequencing coverage, the prevalence of detectable M-CHIP mutations in our WTC cohort was 7.5% (26/345) and L-CHIP was 9.9% (34/345), while in unexposed controls M-CHIP was 3.1% (9/293) and L-CHIP was 2.0% (6/293). Notably, L-CHIP mutations were statistically significantly more prevalent in WTC participants versus controls (*p*=0.031). However, M-CHIP prevalence did not reach statistical significance (*p* = 0.693). At gene level, *EEF1A1* (*p*=0.024) and *DDX11* (*p*=0.071) were elevated in WTC responders (Supplementary Figure S4).

This is the first comprehensive characterization of CHIP mutation patterns across occupational groups in WTC responders, leveraging extensive clinical, laboratory, cognitive and HLA zygosity data, and the first characterization of L-CHIP in a WTC-associated cohort. Our findings expand on previous research focused primarily on firefighters^5^, given the documented elevated cancer risks for those who were not firefighters^6^. Furthermore, using deep WES allowed an unbiased survey across myeloid and lymphoid genes, creating a valuable resource for future studies as CHIP definitions evolve.

This study has broader implications. Other populations exposed to large-scale fires and collapses, including civilians in modern warfare in populated cities may also face elevated CHIP risks. Future research should aim to refine cancer risk prediction models that include CHIP status, blood counts, and demographic factors to guide early detection strategies. Although relative risks are increased, the absolute risk of malignancy in CHIP-positive individuals remains low, underscoring the need for careful risk stratification rather than blanket interventions. As CHIP clinics emerge at academic centers, longitudinal data will inform improved risk models, allowing high-risk individuals to be monitored appropriately and potentially enrolled in clinical trials.

Study limitations include occupational confounders in our unexposed controls, inability to detect mosaic chromosomal alterations via WES, modest sample size limiting detection of subtle associations (particularly for L-CHIP), and the risk of false positives from multiple comparisons. Independent validation cohorts will be critical to confirm these findings.

## Data Availability

We will deposit the whole exome sequencing data of the WTC responders in the database of Genotypes and Phenotypes (dbGaP). The CHIP mutation data is contained in the manuscript.

## Code Availability

All analyses were performed utilizing standard publicly available software. Any specific analysis code details are available from the authors upon request.

## Authors’ contributions

P.B and Z.H.G. conceived and designed the study. M.E.S., P.F.K., R.J.K., and Z.H.G. wrote the manuscript. B.J.L. and X.Y. recruited participants and handled sample and data collection. Z.H.G led sample sequencing at Azenta Inc. M.E.S. performed sequence analyses and P.F.K. performed statistical analyses. All authors were involved in the interpretation of the results. B.J.L., J.M. and P.B. edited the manuscript. Z.H.G. supervised the study. All authors approved the final manuscript.

