## supplemental materials for "Distinct characteristics of lymphoid and myeloid clonal hematopoiesis in World Trade Center first responders"

**Conflict of interest.** The authors declare no potential conflicts of interest.

**Ethics statement.** The study received annual approval under IRB #604113 from the Committees on Research Involving Human Subjects at SBU.

**Data availability.** We will deposit the whole exome sequencing data of the WTC responders in the database of Genotypes and Phenotypes (dbGaP).

**Funding.** This work was supported by grants to P.B., J.M. and Z.H.G. from the Centers for Disease Control and Prevention, National Institute of Occupational; Safety and Health (award # 1U01OH012187-01); to Z.H.G from Cancer Moonshot R33 award # CA263705-01; to B.J.L. and X.Y. from the Centers for Disease Control and Prevention (CDC/NIOSH 75D301-22-C-15522); and in part through the computational and data resources and staff expertise provided by Scientific Computing and Data at the Icahn School of Medicine at Mount Sinai and supported by the Clinical and Translational Science Awards (CTSA) grant UL1TR004419 from the National Center for Advancing Translational Sciences.

### Supplemental Methods

Study Population. We studied participants from the World Trade Center Health Program (WTCHP) General Responder Cohort (GRC), which includes individuals involved in rescue, recovery, and cleanup efforts at the WTC site after 9/11/2001. Eligibility was based on task type, site location, and dates/hours worked or volunteered[1,2]. Our study focused on responders receiving care at the WTC Clinical Center at Stony Brook University (WTC-SBU, B. Luft, PI), established in 2002 to monitor and treat WTC-related conditions in individuals with documented WTC-response experience. The WTC-SBU cohort has been well-characterized in previous studies examining various health conditions associated with response to the WTC disaster, including post-traumatic stress disorder (PTSD), prostate cancer, cognitive impairment, and COVID-19[3–6]. Compared to the broader WTCHP GRC, the WTC-SBU cohort includes relatively more law enforcement personnel and men, and fewer individuals without a high-school degree[4]. All participants underwent routine monitoring visits every 12–18 months, including self-administered physical and mental health questionnaires followed by a physical examination, laboratory tests, spirometry, and a chest radiograph. The study received annual approval under IRB #604113 from the Committees on Research Involving Human Subjects at SBU, with over 95% of participants providing written informed consent for research use of their data.

Sample Collection. Whole blood samples were collected from consented participants during annual clinical checkups between 2016 and January 2019 using Vacutainer Plastic K2EDTA (containing ethylenediaminetetraacetic acid) Tubes (BD, 367527) and stored at -80C until analysis.

Demographics. From the WTC-SBU cohort, we randomly selected 350 samples from 345 unique participants for whole-exome sequencing (WES). The cohort was predominantly male (92.5%) and white (91%), with an age distribution of less than 8% aged $\leq$ 55 years, 49.6% aged 56-60 years, 24.3% aged 61-65 years, 9.3% aged 66-70 years, and 9% older than 70. Regarding smoking status, 4.3% were current smokers, 44.6% were former smokers, and 51% were never-smokers. Detailed participant characteristics are presented in Table 1.

Clinical Data. We collected comprehensive clinical data at each monitoring visit, which include: body mass index (BMI), and mental, cognitive and general health characteristics (including cholesterol and triglyceride levels). WTC exposure history was assessed during enrollment interview at the WTC Health Program and has been described in detail[7]. Briefly, an exposure variable was created using total time spent working at Ground Zero or on the debris pile[7]. Post-traumatic stress disorder (PTSD) symptoms were assessed using the PTSD Checklist (PCL), a 20-item self-report measure modified to assess symptoms over the past month, which has excellent psychometric properties, convergent validity and internal consistency[8]. Mild cognitive impairment (MCI) was measured using the Montreal Cognitive Assessment (MoCA), a widely used objective multidomain test[9].

Blood and Lipid Markers. We measured blood and lipid markers concurrently with blood collection for CHIP analysis from 294 (out of 345) participants. Blood markers include platelet count, basophil count, lymphocyte count, neutrophil count, segmented neutrophil count, eosinophil count, monocyte count, LMR (ratio lymphocyte/monocyte), white blood cell count (WBC), red blood cell count (RBC), mean corpuscular volume (MCV), mean corpuscular hemoglobin (MCH), MCH concentration (MCHC), and red cell distribution width (RDW). Lipid markers include total cholesterol, triglycerides, HDL cholesterol, LDL cholesterol, and very low-density LDL (VLDL).

Deep Whole-Exome Sequencing. Deep WES was performed at Azenta Inc (Burlington, MA, USA), using the HiSeq 2500 System (Illumina, San Diego, CA, USA) following standard protocols. To ensure high sensitivity for detecting CHIP mutations even at low clonal frequencies, we achieved a median coverage of 250X. Sequencing generated 150 bp paired-end read data in standard FASTQ format.

Data Pre-Processing for Variant Discovery. To pre-process and QC raw sequence reads, we used fastp[10]. Specifically, we trimmed the adapters and filtered out bad reads (low quality, too short or too many unknown bases). We then followed the Genome Analysis Toolkit (GATK https://software.broadinstitute.org/gatk) best practices for data pre-processing to generate analysis-ready bam files from the fastq files. Briefly, sequence reads were aligned to the Genome Reference Consortium Human Build GrCh38 using BWA-MEM[11] followed by duplicate marking using Picard (http://broadinstitute.github.io/picard) and base quality score recalibration using the GATK, producing analysis-ready BAM files.

Germline Variant Calling and Kinship Analysis. We called germline variants using HaplotypeCaller in GATK in GVCF mode, jointly genotyped individual gVCF files for all autosomes, filtered variants by variant quality score recalibration in GATK and finally removed sites with ≥ 20% missing data. To exclude genetic duplicates among WTC participants, we performed kinship analysis with germline variants using KING software[12] and identified three duplicate pairs (kinship coefficient > 0.354). One pair represented the same individual sampled at different time points. From these six samples, we included only the most recent sample collected from each participant, leaving 345 unique samples for analysis.

Somatic Variant Calling. After kinship analysis, we performed somatic variant calling on the remaining 345 BAM files using the GATK Mutect2[13] pipeline in tumor-only mode. To exclude likely germline calls and sequencing artifacts, we provided Mutect2 with external reference of germline variants from gnomAD and a Panel Of Normals (PON). For the PON, we used WES data from 70 young individuals (aged ≤ 40 years) from the publicly available Genotype-Tissue Expression (GTEx) cohort (phs000424), who would be less likely to harbor CHIP. High-confidence somatic mutations were identified by applying Mutect2's orientation bias and PASS filters.

Somatic Variant Filtering to identify CHIP carriers. We considered 76 somatic driver genes for M-CHIP[14,15], and 235 genes for L-CHIP[16] (Supplementary Table S1), filtering for predefined CHIP variants with a Mutect2 VAF >2%. For L-CHIP, we specifically filtered for *pathogenic* variants (curated from cBioPortal) or *putative* variants (that alter canonical protein sequence)[16].

To remove likely artifacts, we applied stringent QC filters. Briefly, we filtered for somatic variants at a minimum depth of 20 reads, minimum 3 reads (supporting the mutant allele) and at least one read in both forward and reverse directions (supporting the reference and mutant alleles). In addition, we excluded somatic variants observed in gnomAD with allele frequency ≥ 0.1% and with an observed frequency >1% in the cohort (unless previously reported to be involved in hematologic malignancies). Variants were annotated for pathogenicity using for Combined Annotation Dependent Depletion (CADD)[17] score, excluding those with scaled CADD score < 10. For *putative* L-CHIP mutations, we implemented additional filters (Supplementary Table S1), including a minimum alternate allele read of 5, maximum VAF of 0.2 and at least two reads in both forward and reverse directions supporting the alternate allele.

We made two specific exceptions to our filtering protocol: For the *U2AF1* gene, known to have problematic alignment in GrCh38 reference genome due to unintended replication of the *U2AF1* locus in chromosome 21[18], we realigned *U2AF1* using build GrCh37 and repeated somatic variant calling. Second, we recognized that the *ASXL1*- G646Wfs*12 variant with VAF ≥ 10% represents a true CHIP variant[19] rather than a sequencing artifact. However, we did not identify any CHIP mutations in *U2AF1* gene or at *ASXL1*-G646Wfs*12 in our cohort. After filtering, all identified M/L-CHIP mutations are listed in Supplementary Table S3.

<u>Comparative Analysis with Unexposed Controls.</u> To compare CHIP prevalence in WTC responders versus unexposed controls, we used CHIP calls from 293 healthy controls from the Mount Sinai Crohn's and Colitis Registry (MSCCR) cohort (STUDY-11-01669) processed using identical analytical pipelines as above[20,21]. Control cohort characteristics are provided in Supplementary Table S2. To ensure comparable median sequencing coverage, we down-sampled the WTC data using Picard's DownsampleSam tool to 41.5% of total reads, matching the total number of aligned bases (PF_ALIGNED_BASES) between cohorts.

<u>HLA Zygosity.</u> To examine potential associations between zygosity of HLA alleles and CHIP prevalence, we performed HLA typing using HLA-HD[22], determining HLA class I and class II alleles for each participant with precision up to 6-digits.

<u>Statistical Analysis.</u> We used standard statistical methods to understand the association of CHIP with all available factors. Categorical variables were summarized as counts and percentages, and analyzed using Fisher's exact tests. Continuous variables were summarized using median and median absolute deviation, and analyzed using Wilcoxon rank sum tests. For association analyses, as the majority of the responders were White, we collapsed race into two categories: White and non-White/ unknown. We collapsed WTC exposure into three categories, including very low/low, intermediate and high/very high. Missing values were excluded from the analyses.

We defined several WTC outcome subgroups: a) CHIP positive (at least one M or L-CHIP mutation, N=118), b) M-CHIP positive, (at least one M-CHIP mutation, N=56), c) L-CHIP positive (at least one L-CHIP mutation, N=74), d) *DNMT3A* mutation (at least one mutation on *DNMT3A* gene, N=22), e) *TET2* mutation (at least one mutation in *TET2* gene, N=15), f) *PPM1D* mutation (at least one mutation in *PPM1D* gene, N=11), g) *EEF1A1* mutation (at least one mutation in *EEF1A1* gene, N=18), and h) *DDX11* mutation (at least one mutation in *DDX11* gene, N=13). For all analyses participants without any M/ L-CHIP mutations were classified as CHIP-negative (N=227).

We first performed univariate analysis using individual factors as covariates, with false discovery rate adjusted *p*-values reported in Supplementary Table S4. For characteristics that showing significant from marginal associations, we fitted multivariate logistic regression models using CHIP presence as the outcome variable, with significant characteristics as covariates. Statistical significance was defined as $p < 0.05$.

To compare the WTC responders and controls, we applied Firth's penalized logistic regression, adjusting for age, race, gender, and smoking status. Consistent with the above association analysis, race was categorized as White/European versus all others; smoking status was defined as former/current smoker versus never smoker; and age was treated as a continuous variable in the model. Statistical significance was defined as $p < 0.05$.

**Supplementary Tables**

**Supplementary Table S1.** List of (**A**) myeloid associated (M-CHIP) mutations and (**B**) lymphoid associated (L-CHIP) mutations considered in this study.

**Supplementary Table S2.** Characteristics of the 293 unexposed controls from the MSCCR cohort.

**Supplementary Table S3.** All CHIP mutations identified among the 345 WTC samples. (**A**) M-CHIP mutations identified in WTC samples; (**B**) L-CHIP mutations identified in WTC samples; (**C**) Distribution of M-CHIP mutations by genes; (**D**) Distribution of L-CHIP mutations by genes; (**E**) M-CHIP mutations identified in downsampled WTC samples; (**F**) L-CHIP mutations identified in downsampled WTC samples; (**G**) M-CHIP mutations identified in controls; (**H**) L-CHIP mutations identified in controls.

**Supplementary Table S4**. CHIP-phenotype associations. (**A**) CHIP; (**B**) M-CHIP; (**C**) L-CHIP; (**D**) *DNMT3A*; (**E**) *TET2*; (**F**) *PPM1D*; (**G**) *EEF1A1*; (**H**) *DDX11*.

**Supplementary Table S5.** Prevalence of M-CHIP and L-CHIP mutations in down-sampled 345 WTC debris-exposed first responders and 293 unexposed controls.

**Supplementary Table S2.** Characteristics of the 293 unexposed controls from the MSCCR cohort.

| Phenotype | Category | # Samples (%) |
|---|---|---|
| Sex (M/F =1.05) | Male (M) | 150 (51.2%) |
| | Female (F) | 143 (48.8%) |
| Smoking status | Smoker | 93 (31.7%) |
| | Never smoker | 200 (68.3%) |
| Race | European | 151 (51.5%) |
| | African-American | 65 (22.2%) |
| | AMI | 68 (23.2%) |
| | East Asian | 9 (3.1%) |
| Age (yrs)<br>Median= 54<br>SD= 11.20 | ≤ 55 | 160 (54.6%) |
| | 56 - 60 | 67 (22.9%) |
| | 61 - 65 | 40 (13.7%) |
| | > 65 | 26 (8.9%) |

**Supplementary Table S5.** Prevalence of M-CHIP and L-CHIP mutations in down-sampled 345 WTC debris-exposed first responders and 293 unexposed controls.

| Age (Years) | # Samples | | M-CHIP mutations | | L-CHIP mutations | |
|---|---|---|---|---|---|---|
| | WTC | Control | # WTC | # Control | # WTC | # Control |
| ≤ 55 | 27 | 160 | 0 (0.0%) | 1 (0.6%) | 3 (11.1%) | 3 (1.9%) |
| 56-60 | 171 | 67 | 5 (2.9%) | 3 (4.5%) | 13 (7.6%) | 1 (1.5%) |
| 61-65 | 84 | 40 | 9 (10.7%) | 2 (5.0%) | 9 (10.7%) | 1 (2.5%) |
| > 65 | 63 | 26 | 12 (19.0%) | 3 (11.5%) | 9 (14.3%) | 1 (3.8%) |

### Supplementary Figures

**Supplementary Figure S1. Characteristics of M-CHIP and L-CHIP mutations in 345 WTC debris-exposed first-responders. A)** Number of participants with 1, 2, 3 and 4 M-CHIP mutations; **B)** Number of different types of M-CHIP mutations; **C)** Number of participants with 1,2, 3 and 4 L-CHIP mutations; **D)** Number of different types of L-CHIP mutations.

**Supplementary Figure S2. Variant allele fractions (VAFs) of M-CHIP and L-CHIP mutations. A)** VAF histogram of M-CHIP mutations; **B)** VAF histogram of L-CHIP mutations; **C)** VAF distribution of top M-CHIP gene mutations; **D)** VAF distribution of top L-CHIP gene mutations. Based on VAF, *TET2* exhibited the largest M-CHIP clones, while *EEF1A1* and *DDX11* exhibited the largest L-CHIP clones.

**Supplementary Figure S3. Comparison of key laboratory parameters between different groups. A)** Absolute lymphocyte counts; **B)** Segmented neutrophil counts; **C)** Platelet counts; **D)** Red blood cell (RBC) counts; **E)** Mean corpuscular hemoglobin (MCH); **F)** Mean corpuscular volume (MCV) **G)** Lymphocyte-to-monocyte ratio (LMR)**.**

**Supplementary Figure S4. Characteristics of M-CHIP and L-CHIP mutations in 345 WTC debris-exposed individuals (blue) and 293 unexposed controls (red). A)** Genes with M-CHIP mutations; **B)** Genes with L-CHIP mutations.

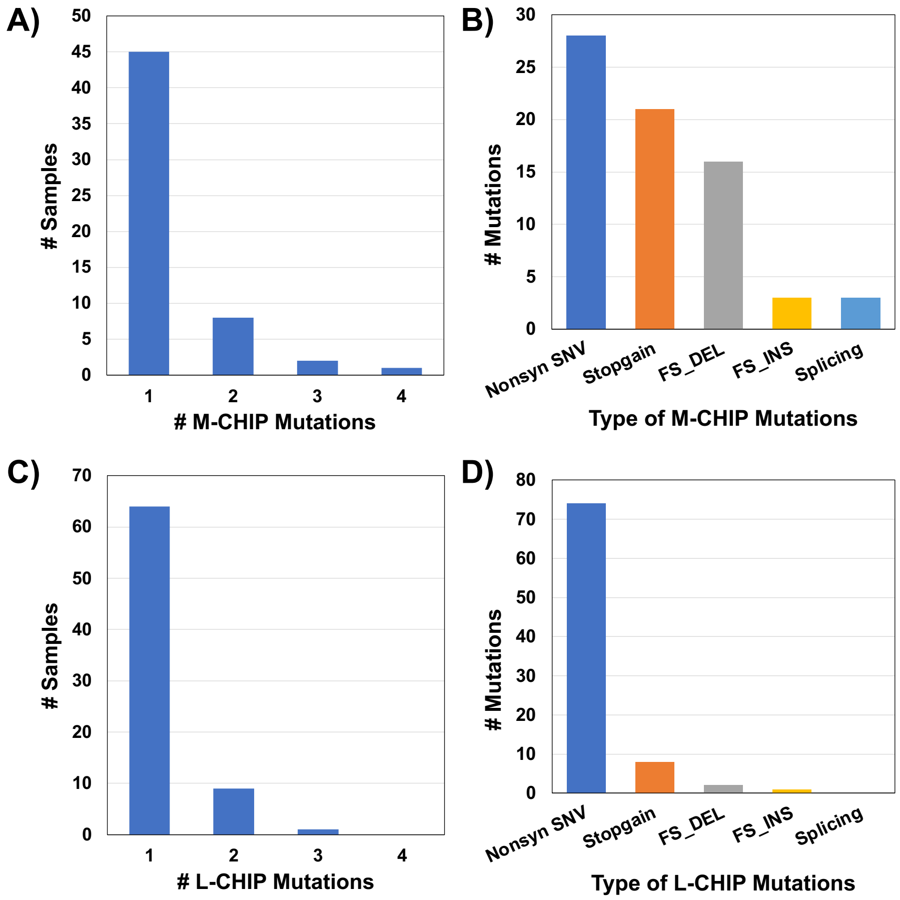


**Supplementary Figure S1. Characteristics of M-CHIP and L-CHIP mutations in 345 WTC debris-exposed first-responders. A)** Number of participants with 1, 2, 3 and 4 M-CHIP mutations, **B)** Number of different types of M-CHIP mutations, **C)** Number of participants with 1,2, 3 and 4 L-CHIP mutations, **D)** Number of different types of L-CHIP mutations.

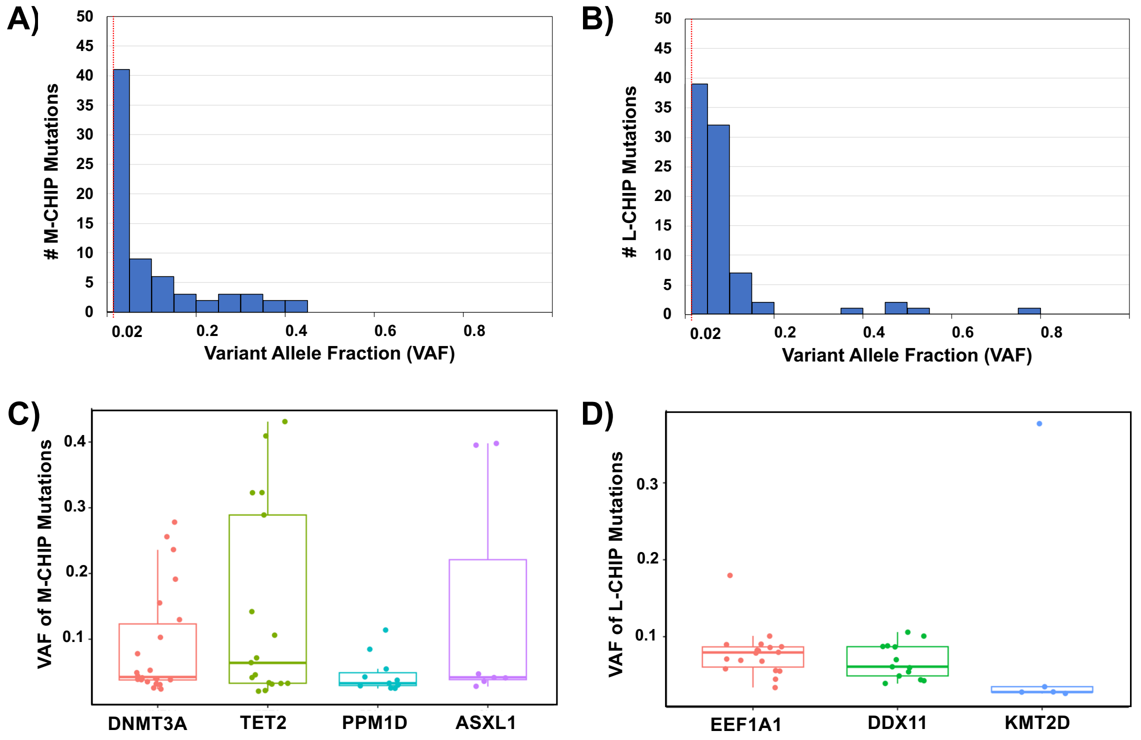


**Supplementary Figure S2. Variant allele fractions (VAFs) of M-CHIP and L-CHIP mutations. A)** VAF histogram of M-CHIP mutations; **B)** VAF histogram of L-CHIP mutations; **C)** VAF distribution of top M-CHIP gene mutations; **D)** VAF distribution of top L-CHIP gene mutations. Based on VAF, *TET2* exhibited the largest M-CHIP clones, while *EEF1A1* and *DDX11* exhibited the largest L-CHIP clones.

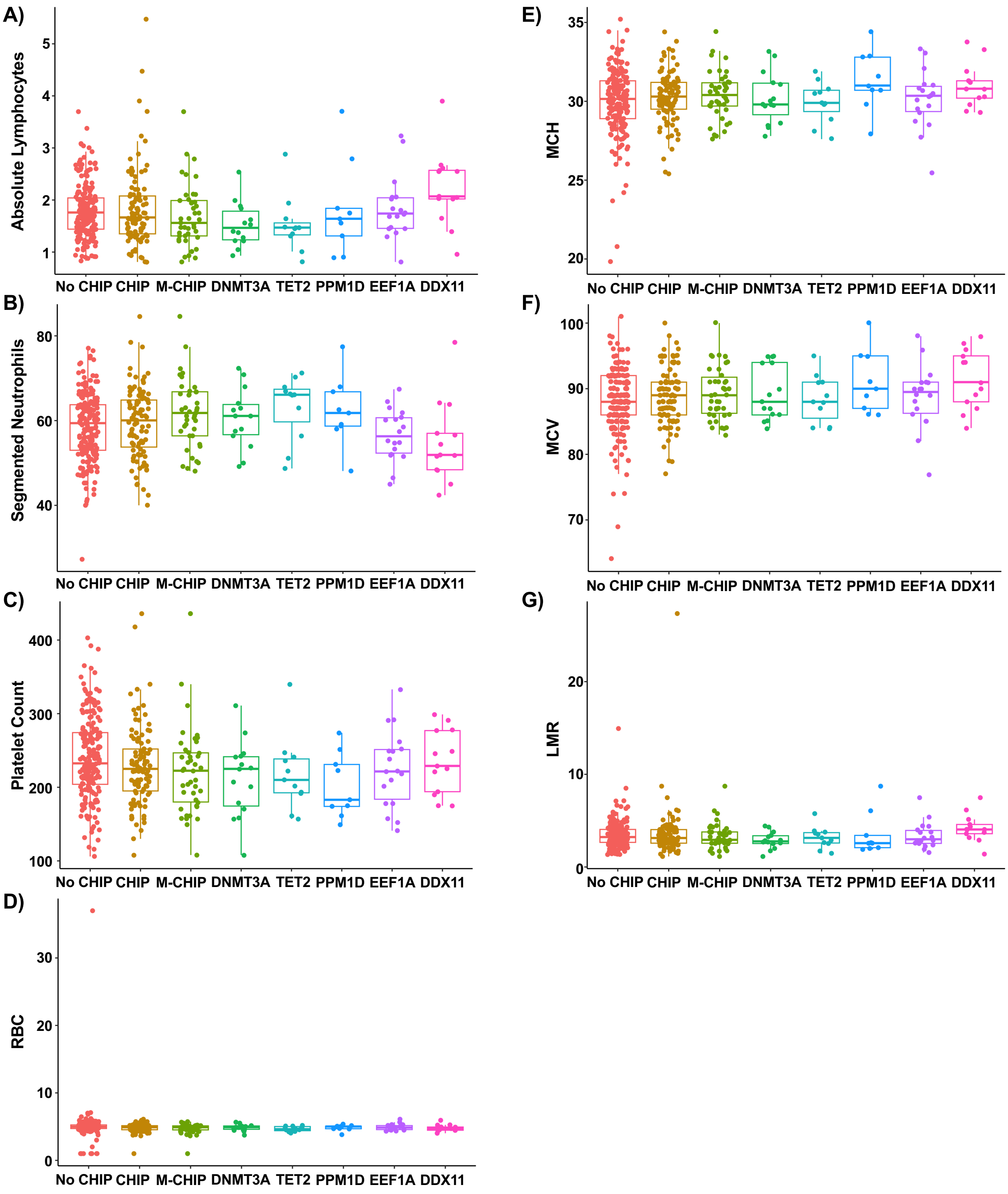


**Supplementary Figure S3. Comparison of key laboratory parameters between different groups. A)** Absolute lymphocyte counts; **B)** Segmented neutrophil counts; **C)** Platelet counts; **D)** Red blood cell (RBC) counts; **E)** Mean corpuscular hemoglobin (MCH); **F)** Mean corpuscular volume (MCV); **G)** Lymphocyte-to-monocyte ratio (LMR)**.**

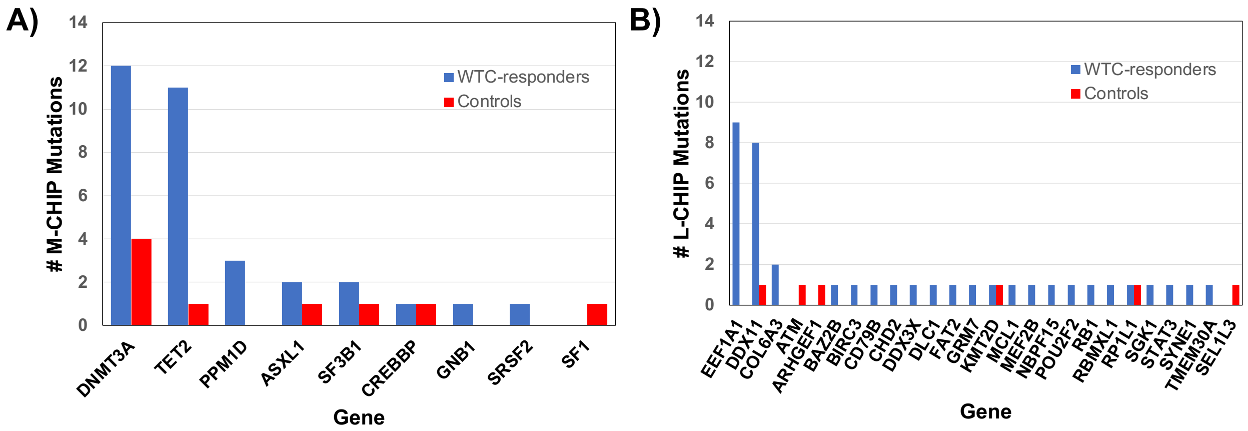


**Supplementary Figure S4. Characteristics of M-CHIP and L-CHIP mutations in 345 WTC debris-exposed responders (blue) and 293 unexposed controls (red). A)** Genes with M-CHIP mutations; **B)** Genes with L-CHIP mutations.
