## Supplementary Table S1 for "Distinct characteristics of lymphoid and myeloid clonal hematopoiesis in World Trade Center first responders"

**Supplementary Table S1a: Myeloid associated M-CHIP Mutations**

| Gene | Mutations | Accession number |
|---|---|---|
| ASXL1 | Frameshift/nonsense/splice-site in exon 11-12 | NM_015338 |
| ASXL2 | Frameshift/nonsense/splice-site in exon 11-12 | NM_018263 |
| BCOR | Frameshift/nonsense/splice-site | NM_001123385 |
| BCORL1 | Frameshift/nonsense/splice-site | NM_021946 |
| BRAF | G464E, G464V, G466E, G466V, G469R, G469E, G469A, G469V, V471F, V472S, L485W, N581S, I582M, I592M, I592V, D594N, D594G, D594V, D594E, F595L, F595S, G596R, L597V, L597S, L597Q, L597R, A598V, V600M, V600L, V600K, V600R, V600E, V600A, V600G, V600D, K601E, K601N, R603*, W604R, W604G, S605G, S605F, S605N, G606E, G606A, G606V, H608R, H608L, G615R, S616P, S616F, L618S, L618W | NM_004333 |
| BRCC3 | Frameshift/nonsense/splice-site | NM_024332 |
| CALR | Frameshift (C-terminal) AA352-418 (exon 9) | NM_004343 |
| CBL | RING finger missense p.381-421 | NM_005188 |
| CBLB | RING finger missense p.372-412 | NM_170662 |
| CEBPA | Frameshift/nonsense/splice-site | NM_004364 |
| CREBBP | Frameshift/nonsense/splice-site, D1435E, R1446L, R1446H, R1446C, Y1450C, P1476R, Y1482H, H1487Y, W1502C, Y1503D, Y1503H, Y1503F, S1680del | NM_004380 |
| CSF1R | L301F, L301S, Y969C, Y969N, Y969F, Y969H, Y969D | NM_005211 |
| CSF3R | T615A, T618I, truncating c.741-791 | NM_000760 |
| CTCF | Frameshift/nonsense, R377C, R377H, P378A, P378L | NM_006565 |
| CUX1 | Frameshift/nonsense | NM_181552 |
| DNMT3A | Frameshift/nonsense/splice-site, F290I, F290C, V296M, P307S, P307R, R326H, R326L, R326C, R326S, G332R, G332E, V339A, V339M, V339G, L344Q, L344P, R366P, R366H, R366G, A368T, A368V, R379H, R379C, I407T, I407N, I407S, F414L, F414S, F414C, A462V, K468R, C497G, C497Y, Q527H, Q527P, Y533C, S535F, C537G, C537R, G543A, G543S, G543C, L547H, L547P, L547F, M548I, M548K, G550R, W581R, W581G, W581C, R604Q, R604W, R635W, R635Q, S638F, G646V, G646E, L653W, L653F, I655N, V657A, V657M, R659H, Y660C, V665G, V665L, M674V, R676W, R676Q, G685R, G685E, G685A, D686Y, D686G, R688H, G699R, G699S, G699D, P700L, P700S, P700R, P700Q, P700T, P700A, D702N, D702Y, V704M, V704G, I705F, I705T, I705S, I705N, G707D, G707V, C710S, C710Y, S714C, V716D, V716F, V716I, N717S, N717I, P718L, R720H, R720G, K721R, K721T, Y724C, R729Q, R729W, R729G, F731C, F731L, F731Y, F731I, F732del, F732C, F732S, F732L, E733G, E733A, F734L, F734C, Y735C, Y735N, Y735S, R736H, R736C, R736P, L737H, L737V, L737F, L737R, A741V, P742P, P743R, P743L, R749C, R749L, R749H, R749G, F751L, F751C, F752del, F752C, F752L, F752I, F752V, W753G, W753C, W753R, L754P, L754R, L754H, F755S, F755I, F755L, M761I, M761V, G762C, V763I, S770L, S770W, S770P, R771Q, F772I, F772V, L773R, L773V, E774K, E774D, E774G, I780T, D781G, R792H, W795C, W795L, G796D, G796V, N797Y, N797H, N797S, P799S, P799R, P799H, R803S, R803W, P804L, P804S, K826R, S828N, K829R, T835M, N838D, K841Q, Q842E, P849L, D857N, W860R, E863D, F868S, G869S, G869V, M880V, S881R, S881I, R882H, R882P, R882C, R882G, A884P, A884V, Q886R, L889P, L889R, G890D, G890R, G890S, V895M, P896L, V897G, V897D, R899L, R899H, R899C, L901R, L901H, P904L, F909C, P904Q, A910P, C911R, C911Y | NM_022552 |
| EED | Frameshift/nonsense/splice-site, L240Q, I363M | NM_003797 |
| EP300 | Frameshift/nonsense/splice_site, VF1148_1149del, D1399N, D1399Y, P1452L, Y1467N, Y1467H, Y1467C, R1627W, A1629V | NM_001429 |
| ETNK1 | N244S, N244T, N244K | NM_018638 |
| ETV6 | Frameshift/nonsense/splice-site | NM_001987 |
| EZH2 | Frameshift/nonsense/splice-site, Q62R, N102S, F145S, F145C, F145Y, F145L, G159R, E164D, R202Q, K238E, E244K, R283Q, H292R, P488S, R497Q, R561H, T568I, K629E, Y641N, Y641H, Y641S, Y641C, Y641F, D659Y, D659G, V674M, A677G, A677V, R679C, R679H, R685C, R685H, A687V, N688I, N688K, H689Y, S690P, I708V, I708T, I708M, E720K, E740K | NM_001203247 |
| FLT3 | V579A, V592A, V592I, F594L, FY590-591GD, D835Y, D835H, D835E, del835 | NM_004119 |
| GATA1 | Frameshift/nonsense/splice-site | NM_002049 |
| GATA2 | Frameshift/nonsense/splice-site, R293Q, N317H, A318T, A318V, A318G, G320D, L321P, L321F, L321V, Q328P, R330Q, R361L, L359V, A372T, R384G, R384K | NM_001145661 |
| GATA3 | Frameshift/nonsense/splice-site ZNF domain, R276W, R276Q, N286T, L348V, | NM_001002295 |
| GNA13 | I34T, G57S, S62F, M68K, Q134R, Y145F, L152F, E167D, Q169H, R264H, E273K, V322G, V362G, L371F | NM_006572 |
| GNAS | R201S, R201C, R201H, R201L, Q227K, Q227R, Q227L, Q227H, R374C, R844S, R844C, R844H, R844L, Q870K, Q870R, Q870L, Q870H, R1017C | NM_000516 |
| GNB1 | K57N, K57M, K57E, K57T, I80T, I80N | NM_002074 |
| IDH1 | R132C, R132G, R132H, R132L, R132P, R132V, V178I | NM_005896 |
| IDH2 | R140W, R140Q, R140L, R140G, R172W, R172G, R172K, R172T, R172M, R172N, R172S | NM_002168 |
| IKZF1 | Frameshift/nonsense | NM_006060 |
| IKZF2 | Frameshift/nonsense | NM_016260 |

| | | |
|---|---|---|
| IKZF3 | Frameshift/nonsense | NM_012481 |
| JAK1 | T478A, T478S, V623A, A634D, L653F, R724H, R724Q, R724P, T782M, L783F | NM_002227 |
| JAK2 | N533D, N533Y, N533S, H538R, K539E, K539L, I540T, I540V, V617F, R683S, R683G, del/ins537-539L, del/ins538-539L, del/ins540-543MK, del/ins540-544MK, del/ins541-543K, del542-543, del543-544, ins11546-547 | NM_004972 |
| JAK3 | M511T, M511I, A572V, A572T, A573V, R657Q, V715I, V715A | NM_000215 |
| KDM6A | Frameshift/nonsense/splice-site, del419 | NM_021140 |
| KIT | ins503, V559A, V559D, V559G, V559I, V560D, V560A, V560G, V560E, del560, E561K, del579, P627L, P627T, R634W, K642E, K642Q, V654A, V654E, H697Y, H697D, E761D, K807R, D816H, D816Y, D816F, D816I, D816V, D816H, del551-559 | NM_000222 |
| KRAS | G12D, G12A, G12E, G12V, G13D, G13C, G13Y, G13F, G13R, G13A, G13V, G13E, V14I, T58I, G60D, G60A, G60V, Q61K, Q61E, Q61P, Q61R, Q61L, Q61H, K117E, K117N, A146T, A146P, A146V | NM_033360 |
| LUC7L2 | Frameshift/nonsense/splice-site | NM_016019 |
| MLL | Frameshift/nonsense | NM_005933 |
| MLL2 | Frameshift/nonsense | NM_003482 |
| MPL | S505G, S505N, S505C, L510P, del513, W515A, W515R, W515K, W515S, W515L, A519T, A519V, Y591D, W515-518KT | NM_005373 |
| NF1 | Frameshift/nonsense | NM_000267 |
| NPM1 | Frameshift p.W288fs (insertion at c.859_860, 860_861, 862_863, 863_864) | NM_002520 |
| NRAS | G12S, G12R, G12C, G12N, G12P, G12Y, G12D, G12A, G12V, G12E, G13S, G13R, G13C, G13N, G13P, G13Y, G13D, G13A, G13V, G13E, G60E, G60R, Q61R, Q61L, Q61K, Q61P, Q61H, Q61Q | NM_002524 |
| PDS5B | Frameshift/nonsense/splice-site, R1292Q | NM_015032 |
| PDSS2 | Frameshift/nonsense | NM_020381 |
| PHF6 | Frameshift/nonsense/splice-site, A40D, M125I, S246Y, F263L, R274Q, C297Y, H302Y, H329L | NM_001015877 |
| PHIP | Frameshift/nonsense/splice-site | NM_017934 |
| PIGA | Frameshift/nonsense/splice-site | NM_002641 |
| PPM1D | Frameshift/nonsense, exon 5 or 6 | NM_003620 |
| PRPF40B | Frameshift/nonsense/splice-site, P15H, M58I, P405L, P562S, | NM_001031698 |
| PRPF8 | M1307I, C1594W, D1598Y, D1598N, D1598V | NM_006445 |
| PTEN | Frameshift/nonsense/splice-site, D24G, R47G, F56V, L57W, H61R, K66N, Y68H, C71Y, F81C, Y88C, D92G, D92V, D92E, H93Y, H93D, H93Q, N94I, P95L, I101T, C105F, C105S, D107Y, L112V, H123Y, C124R, C124S, K125E, A126D, K128N, R130G, R130Q, R130L, G132D, I135V, I135K, C136R, C136F, K144Q, A151T, D153Y, D153N, Y155H, Y155C, R159K, R159S, R161K, R161I, G165R, G165E, S170N, S170I, R173C, Y174D, Y177C, H196Y, R234W, G251C, D252Y, F271S, D326G | NM_000314 |
| PTPN11 | G60V, G60R, G60A, D61Y, D61V, D61G, Y63C, E69K, E69G, E69D, E69Q, F71L, F71K, A72T, A72V, A72D, T73I, E76K, E76Q, E76M, E76A, E76G, E139G, E139D, N308D, N308T, N339S, P491L, S502P, S502A, S502L, G503V, G503G, G503A, G503E, Q506P, T507A, T507K | NM_002834 |
| RAD21 | Frameshift/nonsense/splice-site, R65Q, H208R, Q474R | NM_006265 |
| RUNX1 | Frameshift/nonsense/splice-site, S73F, H78Q, H78L, R80C, R80P, R80H, L85Q, P86L, P86H, S114L, D133Y, L134P, R135G, R135K, R135S, R139Q, R142S, A165V, R174Q, R177L, R177Q, A224T, D171G, D171V, D171N, R205W, R223C | NM_001001890 |
| SETBP1 | D868N, D868T, S869N, G870S, I871T, D880N, D880Q | NM_015559 |
| SETD2 | Frameshift/nonsense, V1190M | NM_014159 |
| SETDB1 | Frameshift/nonsense, K715E | NM_001145415 |
| SF1 | Frameshift/nonsense/splice-site, T454M, Y476C, A508G | NM_004630 |
| SF3A1 | Frameshift/nonsense/splice-site, A57S, M117I, K166T, Y271C | NM_005877 |
| SF3B1 | G347V, R387W, R387Q, E592K, E622D, Y623C, R625L, R625C, R625G, H662Q, H662D, T663I, K666N, K666T, K666E, K666R, K700E, V701F, A708T, G740R, G740E, A744P, D781G, E783K, R831Q, L833F, E862K, R957Q | NM_012433 |
| SRSF2 | Y44H, P95H, P95L, P95T, P95R, P95A, P107H, P95fs | NM_003016 |
| SMC1A | K190T, R586W, M689V, R807H, R1090H, R1090C | NM_006306 |
| SMC3 | Frameshift/nonsense, R155I, Q367E, D392V, K571R, R661P, G662C | NM_005445 |
| STAG1 | Frameshift/nonsense/splice-site, H1085Y | NM_005862 |
| STAG2 | Frameshift/nonsense/splice-site | NM_006603 |
| SUZ12 | Frameshift/nonsense | NM_015355 |
| TET2 | Frameshift/nonsense/splice-site, missense mutations in catalytic domains (p.1104-1481 and 1843-2002) | NM_001127208 |

| | | |
|---|---|---|
| TP53 | Frameshift/nonsense/splice-site, S46F, G105C, G105R, G105D, G108S, G108C, R110L, R110C, T118A, T118R, T118I, S127F, S127Y, L130V, L130F, K132Q, K132E, K132W, K132R, K132M, K132N, F134V. F134L, F134S, C135W, C135S, C135F, C135G, C135Y, Q136K, Q136E, Q136P, Q136R, Q136L, Q136H, A138P, A138V, A138A, A138T, T140I, C141R, C141G, C141A, C141Y, C141S, C141F, C141W, V143M, V143A, V143E, L145Q, W146C, W146L, L145R, V147G, P151T, P151A, P151S, P151H, P151R, P152S, P152R, P152L, T155P, T155A, V157F, R158H, R158L, A159V, A159P, A159S, A159D, A161T, A161D, Y163N, Y163H, Y163D, Y163S, Y163C, K164E, K164M, K164N, K164P, H168Y, H168P, H168R, H168L, H168Q, M169I, M169T, M169V, E171K, E171Q, E171G, E171A, E171V, E171D, V172D, V173M, V173L, V173G, R174W, R175G, R175C, R175H, C176R, C176G, C176Y, C176F, C176S, P177R, P177R, P177L, H178D, H178P, H178Q, H179Y, H179R, H179Q, R181C, R181Y, D186G, G187S, P190L, P190T, H193N, H193P, H193L, H193R, L194F, L194R, I195F, I195N, I195T, R196P, V197L, G199V, Y205N, Y205C, Y205H, D208V, R213Q, R213P, R213L, R213Q, H214D, H214R, S215G, S215I, S215R, V216M, V217G, Y220N, Y220H, Y220S, Y220C, E224D, I232F, I232N, I232T, I232S, Y234N, Y234H, Y234S, Y234C, Y236N, Y236H, Y236C, M237V, M237K, M237I, C238R, C238G, C238Y, C238W, N239T, N239S, S241Y, S241C, S241F, C242G, C242Y, C242S, C242F, G244S, G244C, G244D, G245S, G245R, G245C, G245D, G245A, G245V, G245S, M246V, M246K, M246R, M246I, N247I, R248W, R248G, R248Q, R249G, R249W, R249T, R249M, P250L, I251N, L252P, I254S, I255F, I255N, I255S, L257Q, L257P, E258K, E258Q, D259Y, S261T, G262D, G262V, L265P, G266R, G266E, G266V, R267W, R267Q, R267P, E271K, V272M, V272L, R273S, R273G, R273C, R273H, R273P, R273L, V274F, V274D, V274A, V274G, V274L, C275Y, C275S, C275F, A276P, C277F, C277Y, P278T, P278A, P278S, P278H, P278R, P278L, G279E, R280G, R280K, R280T, R280I, R280S, D281N, D281H, D281Y, D281G, D281E, R282G, R282W, R282Q, R282P, E285K, E285V, E286G, E286V, E286K, K320N, L330R, G334V, R337C, R337L, A347T, L348F, T377P | NM_001126112 |
| U2AF1 | D14G, S34F, S34Y, R35L, R156H, R156Q, Q157R, Q157P | NM_006758 |
| U2AF2 | R18W, Q143L, M144I, L187V, Q190L | NM_007279 |
| WT1 | Frameshift/nonsense/splice-site | NM_024426 |
| ZRSR2 | Frameshift/nonsense, R126P, E133G, C181F, H191Y, I202N, F239V, F239Y, N261Y, C280R, C302R, C326R, H330R, N382K | NM_005089 |

**Supplementary Table S1b: Lymphoid associated L-CHIP Mutations**

| Gene | Pathogenic driver mutations | Putative mutations |
|---|---|---|
| ABL1 | | Frameshift/stop gain/splice site, Nonsynonymous |
| ACTB | | Frameshift/stop gain/splice site, Nonsynonymous |
| ACTG1 | Splice site | |
| ADD2 | | Frameshift/stop gain/splice site, Nonsynonymous |
| ADGRV1 | | Frameshift/stop gain/splice site, Nonsynonymous |
| AKAP6 | | Frameshift/stop gain/splice site, Nonsynonymous |
| ALK | | Frameshift/stop gain/splice site, Nonsynonymous |
| ANKRD50 | | Frameshift/stop gain/splice site, Nonsynonymous |
| AOC2 | | Frameshift/stop gain/splice site, Nonsynonymous |
| ARHGEF1 | | Frameshift/stop gain/splice site, Nonsynonymous |
| ARID1A | Frameshift, nonsense, splice site, AAS G2087R | Nonsynonymous |
| ARID1B | Frameshift, nonsense, splice site | |
| ARID5B | Frameshift, nonsense, splice site | |
| ATM | Frameshift, nonsense, splice site, AAS R337H, R2832H/C, N2875S, I2888T, R3008C/H | Nonsynonymous |
| ATR | Frameshift, nonsense, splice site | |
| AUTS2 | | Frameshift/stop gain/splice site, Nonsynonymous |
| B2M | Frameshift, nonsense, splice site, AAS L7V L12P/Q/R | Nonsynonymous |
| BAZ2A | | Frameshift/stop gain/splice site, Nonsynonymous |
| BAZ2B | | Frameshift/stop gain/splice site, Nonsynonymous |
| BCL10 | Frameshift, nonsense, splice site | |
| BCL11A | | Frameshift/stop gain/splice site, Nonsynonymous |
| BCL2 | AAS K22N/R, G33R/E/A, P95A/S/L/R, G101V, A131V/D/G/T, N172D/S | Nonsynonymous |
| BCL6 | AAS R618C | Nonsynonymous |
| BCL7A | | Frameshift/stop gain/splice site, Nonsynonymous |
| BIRC3 | Frameshift, nonsense, splice site | |
| BRCA1 | | Frameshift/stop gain/splice site, Nonsynonymous |
| BRWD3 | | Frameshift/stop gain/splice site, Nonsynonymous |
| BTG1 | Frameshift, nonsense, splice site | |
| BTG2 | | Frameshift/stop gain/splice site, Nonsynonymous |
| BTK | AAS T316A | Nonsynonymous |
| CARD11 | AAS C49Y/S, E93D/Q, R113Q, F115I/L/V, T117P/N, g123C/D/S, G126D, F130C/I/V, K215E/M/N/T, D230N, S250P, L251P, R337Q, D357E/V, Y361H, D387A, D401N/V, R423W, E626K | Nonsynonymous |
| CCAR1 | | Frameshift/stop gain/splice site, Nonsynonymous |
| CCL4 | | Frameshift/stop gain/splice site, Nonsynonymous |
| CCND1 | AAS S41T, V42E, Y44C/D/F/H/N/S, K46E/I/N, C47R/S, T286A, P287S | Nonsynonymous |
| CCND3 | Frameshift, nonsense, splice site at C-terminal end, AAS T283A/I, P284S/R/A/L/T, D286G, I290K/R/T/M | Nonsynonymous |
| CD19 | | Frameshift/stop gain/splice site, Nonsynonymous |
| CD274 | | Frameshift/stop gain/splice site, Nonsynonymous |
| CD58 | Frameshift, nonsense, splice site | |
| CD70 | | Frameshift/stop gain/splice site, Nonsynonymous |
| CD79A | | Frameshift/stop gain/splice site, Nonsynonymous |
| CD79B | AAS Y196H/S/C/N/D/F | Nonsynonymous |
| CD83 | | Frameshift/stop gain/splice site, Nonsynonymous |
| CDKN2A | Frameshift, nonsense, splice site, AAS R112H | Nonsynonymous |
| CHD2 | | Frameshift/stop gain/splice site, Nonsynonymous |
| CHD8 | | Frameshift/stop gain/splice site, Nonsynonymous |
| CHEK2 | Frameshift, nonsense, splice site | |
| CIITA | Frameshift, nonsense, splice site, T636M | Nonsynonymous |
| CLGN | | Frameshift/stop gain/splice site, Nonsynonymous |
| CNKSR2 | | Frameshift/stop gain/splice site, Nonsynonymous |
| COL6A3 | | Frameshift/stop gain/splice site, Nonsynonymous |
| CSF2RB | | Frameshift/stop gain/splice site, Nonsynonymous |
| CTSS | | Frameshift/stop gain/splice site, Nonsynonymous |
| CXCR4 | Frameshift, nonsense, splice site at C-terminal end | Nonsynonymous |
| DAZAP1 | | Frameshift/stop gain/splice site, Nonsynonymous |
| DDX11 | | Frameshift/stop gain/splice site, Nonsynonymous |
| DDX3X | Frameshift, nonsense, splice site, AAS Y525H, R528H, R534H/C | Nonsynonymous |
| DGKB | | Frameshift/stop gain/splice site, Nonsynonymous |
| DIRAS3 | | Frameshift/stop gain/splice site, Nonsynonymous |
| DLC1 | | Frameshift/stop gain/splice site, Nonsynonymous |
| DMD | | Frameshift/stop gain/splice site, Nonsynonymous |
| DTX1 | Frameshift, nonsense, splice site | |
| DUSP2 | | Frameshift/stop gain/splice site, Nonsynonymous |
| DUSP22 | | Frameshift/stop gain/splice site, Nonsynonymous |
| EBF1 | | Frameshift/stop gain/splice site, Nonsynonymous |
| EEF1A1 | | Frameshift/stop gain/splice site, Nonsynonymous |
| ENPP3 | | Frameshift/stop gain/splice site, Nonsynonymous |
| ENTPD4 | | Frameshift/stop gain/splice site, Nonsynonymous |
| ERBB4 | AAS K1223T | Nonsynonymous |
| ETS1 | | Frameshift/stop gain/splice site, Nonsynonymous |
| FANCE | | Frameshift/stop gain/splice site, Nonsynonymous |
| FAS | Frameshift, nonsense, splice site | |

| | | |
|---|---|---|
| FAT1 | Frameshift, nonsense, splice site, AAS R1627Q, A4419V | Nonsynonymous |
| FAT2 | | Frameshift/stop gain/splice site, Nonsynonymous |
| FAT4 | | Frameshift/stop gain/splice site, Nonsynonymous |
| FBXW7 | Frameshift, nonsense, splice site, AAS R465H/C/L, Y545C | Nonsynonymous |
| FOXC1 | | Frameshift/stop gain/splice site, Nonsynonymous |
| FOXO1 | Frameshift, nonsense, splice site, AAS S22P/W, T24I/A, S205N | Nonsynonymous |
| FYN | | Frameshift/stop gain/splice site, Nonsynonymous |
| GNA13 | | Frameshift/stop gain/splice site, Nonsynonymous |
| GNAI2 | | Frameshift/stop gain/splice site, Nonsynonymous |
| GNE | | Frameshift/stop gain/splice site, Nonsynonymous |
| GPS2 | Frameshift, nonsense, splice site | |
| GRB2 | | Frameshift/stop gain/splice site, Nonsynonymous |
| GRHPR | | Frameshift/stop gain/splice site, Nonsynonymous |
| GRM7 | | Frameshift/stop gain/splice site, Nonsynonymous |
| HAVCR2 | | Frameshift/stop gain/splice site, Nonsynonymous |
| HEATR3 | | Frameshift/stop gain/splice site, Nonsynonymous |
| HIST1H1B | Frameshift, nonsense, splice site | |
| HIST1H1C | AAS A180P | Nonsynonymous |
| HIST1H1D | | Frameshift/stop gain/splice site, Nonsynonymous |
| HIST1H1E | | Frameshift/stop gain/splice site, Nonsynonymous |
| HIST1H2AC | | Frameshift/stop gain/splice site, Nonsynonymous |
| HIST1H2AM | | Frameshift/stop gain/splice site, Nonsynonymous |
| HIST1H2BC | AAS E77G/K, A78P, L103F/I | Nonsynonymous |
| HIST1H2BK | | Frameshift/stop gain/splice site, Nonsynonymous |
| HIST1H3B | | Frameshift/stop gain/splice site, Nonsynonymous |
| HIST2H2BE | | Frameshift/stop gain/splice site, Nonsynonymous |
| HNRNPD | | Frameshift/stop gain/splice site, Nonsynonymous |
| HVCN1 | | Frameshift/stop gain/splice site, Nonsynonymous |
| ID3 | Frameshift, nonsense, splice site, AAS P56S/L/R, L64F/H/R/V | Nonsynonymous |
| IGLL5 | | Frameshift/stop gain/splice site, Nonsynonymous |
| IKZF3 | | Frameshift/stop gain/splice site, Nonsynonymous |
| IL10RA | | Frameshift/stop gain/splice site, Nonsynonymous |
| IL6 | | Frameshift/stop gain/splice site, Nonsynonymous |
| IRF2BP2 | | Frameshift/stop gain/splice site, Nonsynonymous |
| IRF4 | AAS C99R | Nonsynonymous |
| IRF8 | Frameshift, nonsense, splice site at C-terminal end, AAS T80A | Nonsynonymous |
| ITK | | Frameshift/stop gain/splice site, Nonsynonymous |
| ITPKB | | Frameshift/stop gain/splice site, Nonsynonymous |
| ITPR3 | | Frameshift/stop gain/splice site, Nonsynonymous |
| JAK1 | AAS Y652H, L910P, Y1035C | Nonsynonymous |
| JAK3 | AAS 573V, R657Q | Nonsynonymous |
| KIAA1671 | | Frameshift/stop gain/splice site, Nonsynonymous |
| KIR2DL3 | | Frameshift/stop gain/splice site, Nonsynonymous |
| KLF2 | Frameshift, nonsense, splice site | |
| KLHL14 | | Frameshift/stop gain/splice site, Nonsynonymous |
| KLHL21 | | Frameshift/stop gain/splice site, Nonsynonymous |
| KLHL6 | | Frameshift/stop gain/splice site, Nonsynonymous |
| KLHL9 | | Frameshift/stop gain/splice site, Nonsynonymous |
| KMT2C | Frameshift, nonsense, splice site | |
| KMT2D | Frameshift, nonsense, splice site, AAS R5179C, R5432W/Q | Nonsynonymous |
| LRP1B | | Frameshift/stop gain/splice site, Nonsynonymous |
| LTB | | Frameshift/stop gain/splice site, Nonsynonymous |
| LYN | | Frameshift/stop gain/splice site, Nonsynonymous |
| MAGT1 | | Frameshift/stop gain/splice site, Nonsynonymous |
| MAP2K1 | | Frameshift/stop gain/splice site, Nonsynonymous |
| MATN2 | | Frameshift/stop gain/splice site, Nonsynonymous |
| MCL1 | | Frameshift/stop gain/splice site, Nonsynonymous |
| MED11 | | Frameshift/stop gain/splice site, Nonsynonymous |
| MEF2B | AAS K4E, Y69N, E77K, N81K/Y, D83V | Nonsynonymous |
| MGA | Frameshift, nonsense, splice site | |
| MGARP | | Frameshift/stop gain/splice site, Nonsynonymous |
| MPEG1 | | Frameshift/stop gain/splice site, Nonsynonymous |
| MS4A1 | | Frameshift/stop gain/splice site, Nonsynonymous |
| MTOR | AAS W1456G, A1459V, C1483Y, A1971T, T1977K, T1977R, V2006I, S2215F, R2217W, S2231L, I2500F | Nonsynonymous |
| MYC | AAS V7L/A/M, T73I/N/A, P74S/A, S161L | Nonsynonymous |
| MYCBP2 | | Frameshift/stop gain/splice site, Nonsynonymous |
| MYD88 | AAS M232T, S243N, L265P | |
| NBPF14 | | Frameshift/stop gain/splice site, Nonsynonymous |
| NBPF15 | | Frameshift/stop gain/splice site, Nonsynonymous |
| NCOR1 | Frameshift, nonsense, splice site | |
| NCOR2 | | Frameshift/stop gain/splice site, Nonsynonymous |
| NEB | | Frameshift/stop gain/splice site, Nonsynonymous |
| NFKBIA | Frameshift, nonsense, splice site | |
| NFKBIE | | Frameshift/stop gain/splice site, Nonsynonymous |

| | | |
|---|---|---|
| NFKBIZ | | Frameshift/stop gain/splice site, Nonsynonymous |
| NLRP8 | | Frameshift/stop gain/splice site, Nonsynonymous |
| NOL9 | | Frameshift/stop gain/splice site, Nonsynonymous |
| NOTCH1 | Frameshift, nonsense, splice site at C-terminal end | |
| NOTCH2 | Frameshift, nonsense, splice site at C-terminal end | |
| NSD2 | AAS E1099K, T1150A | |
| NTRK1 | | Frameshift/stop gain/splice site, Nonsynonymous |
| NUP214 | | Frameshift/stop gain/splice site, Nonsynonymous |
| OR2M3 | | Frameshift/stop gain/splice site, Nonsynonymous |
| OSBPL10 | | Frameshift/stop gain/splice site, Nonsynonymous |
| P2RY8 | | Frameshift/stop gain/splice site, Nonsynonymous |
| PABPC3 | | Frameshift/stop gain/splice site, Nonsynonymous |
| PAX5 | Frameshift, nonsense, splice site, AAS V26G | Nonsynonymous |
| PCBP1 | | Frameshift/stop gain/splice site, Nonsynonymous |
| PCLO | | Frameshift/stop gain/splice site, Nonsynonymous |
| PDCD1 | | Frameshift/stop gain/splice site, Nonsynonymous |
| PDE4DIP | | Frameshift/stop gain/splice site, Nonsynonymous |
| PDGFRB | | Frameshift/stop gain/splice site, Nonsynonymous |
| PEX14 | | Frameshift/stop gain/splice site, Nonsynonymous |
| PIK3CA | AAS R108H | Nonsynonymous |
| PIK3CD | AAS G124D, C416R, E525K, E1021K | Nonsynonymous |
| PIM1 | AAS L2F, N7K, K24N, G28D/A/S/V, Q37H, H68Y, E79D/K, P81S/A/I/T, S97N/T, L193F/I | Nonsynonymous |
| PIM2 | | Frameshift/stop gain/splice site, Nonsynonymous |
| PLCG2 | | Frameshift/stop gain/splice site, Nonsynonymous |
| PLXNB3 | | Frameshift/stop gain/splice site, Nonsynonymous |
| POT1 | Frameshift, nonsense, splice site | |
| POU2AF1 | | Frameshift/stop gain/splice site, Nonsynonymous |
| POU2F2 | | Frameshift/stop gain/splice site, Nonsynonymous |
| PPP4R3A | | Frameshift/stop gain/splice site, Nonsynonymous |
| PRDM1 | Frameshift, nonsense, splice site | |
| PRKCB | | Frameshift/stop gain/splice site, Nonsynonymous |
| PRUNE2 | | Frameshift/stop gain/splice site, Nonsynonymous |
| PTCH1 | | Frameshift/stop gain/splice site, Nonsynonymous |
| PTEN | Frameshift, nonsense, splice site, AAS S10N, L23S, D24E, Y27N, L42F, V45I, H123Y, G129R, I135T, Y155S, R335Q | Nonsynonymous |
| PTPN6 | | Frameshift/stop gain/splice site, Nonsynonymous |
| PTPRD | Frameshift, nonsense, splice site, AAS P1311S, V1565I, R1674H | Nonsynonymous |
| PTPRK | | Frameshift/stop gain/splice site, Nonsynonymous |
| RB1 | Frameshift, nonsense, splice site, AAS R661W, C706R, S758L | Nonsynonymous |
| RBMXL1 | | Frameshift/stop gain/splice site, Nonsynonymous |
| RCOR1 | | Frameshift/stop gain/splice site, Nonsynonymous |
| REL | | Frameshift/stop gain/splice site, Nonsynonymous |
| RET | | Frameshift/stop gain/splice site, Nonsynonymous |
| RHOA | AAS R5Q/W, G17A, Y34N, N41S, Y42C/F/S/H, Y66N, L69P/R, A161T | Nonsynonymous |
| ROS1 | | Frameshift/stop gain/splice site, Nonsynonymous |
| RP1L1 | | Frameshift/stop gain/splice site, Nonsynonymous |
| RPS15 | | Frameshift/stop gain/splice site, Nonsynonymous |
| SCG3 | | Frameshift/stop gain/splice site, Nonsynonymous |
| SEL1L3 | | Frameshift/stop gain/splice site, Nonsynonymous |
| SETD1B | | Frameshift/stop gain/splice site, Nonsynonymous |
| SETD5 | | Frameshift/stop gain/splice site, Nonsynonymous |
| SETDB1 | | Frameshift/stop gain/splice site, Nonsynonymous |
| SGK1 | | Frameshift/stop gain/splice site, Nonsynonymous |
| SIN3A | | Frameshift/stop gain/splice site, Nonsynonymous |
| SLC9A6 | | Frameshift/stop gain/splice site, Nonsynonymous |
| SMARCA4 | Frameshift, nonsense, splice site, AAS T910M/A, R973Q/L, R1135Q, R1189L, R1192C/H, R1243L/W | Nonsynonymous |
| SOCS1 | Frameshift, nonsense, splice site, AAS A3T/S/P/V | Nonsynonymous |
| SOCS6 | | Frameshift/stop gain/splice site, Nonsynonymous |
| SPEN | Frameshift, nonsense, splice site | |
| SPIB | | Frameshift/stop gain/splice site, Nonsynonymous |
| STAT3 | AAS in SH2 domain (S614N/R, E616G/K, Y640F, K658R, D661V/Y) | Nonsynonymous |
| STAT6 | | Frameshift/stop gain/splice site, Nonsynonymous |
| SYNE1 | | Frameshift/stop gain/splice site, Nonsynonymous |
| TAF1 | | Frameshift/stop gain/splice site, Nonsynonymous |
| TBL1XR1 | AAS D370N, W376C, Y395C/H | Nonsynonymous |
| TCL1A | | Frameshift/stop gain/splice site, Nonsynonymous |
| TCTN2 | | Frameshift/stop gain/splice site, Nonsynonymous |
| TLR2 | | Frameshift/stop gain/splice site, Nonsynonymous |
| TMEM30A | | Frameshift/stop gain/splice site, Nonsynonymous |
| TMSB4X | | Frameshift/stop gain/splice site, Nonsynonymous |
| TNFAIP3 | Frameshift, nonsense, splice site | |
| TNFRSF14 | Frameshift, nonsense, splice site | |
| TNIP1 | | Frameshift/stop gain/splice site, Nonsynonymous |
| TOX | | Frameshift/stop gain/splice site, Nonsynonymous |
| TP63 | | Frameshift/stop gain/splice site, Nonsynonymous |

| | | |
|---|---|---|
| TRAF3 | | Frameshift/stop gain/splice site, Nonsynonymous |
| TSC2 | Frameshift, nonsense, splice site | |
| TUT4 | | Frameshift/stop gain/splice site, Nonsynonymous |
| UBE2A | | Frameshift/stop gain/splice site, Nonsynonymous |
| UBR5 | | Frameshift/stop gain/splice site, Nonsynonymous |
| USP6 | | Frameshift/stop gain/splice site, Nonsynonymous |
| VAV1 | | Frameshift/stop gain/splice site, Nonsynonymous |
| WDFY3 | | Frameshift/stop gain/splice site, Nonsynonymous |
| XPO1 | AAS E571K/V/Q/G | Nonsynonymous |
| YY1 | | Frameshift/stop gain/splice site, Nonsynonymous |
| ZBTB11 | | Frameshift/stop gain/splice site, Nonsynonymous |
| ZC3H12A | | Frameshift/stop gain/splice site, Nonsynonymous |
| ZEB2 | | Frameshift/stop gain/splice site, Nonsynonymous |
| ZFP36L1 | | Frameshift/stop gain/splice site, Nonsynonymous |
| ZNF217 | | Frameshift/stop gain/splice site, Nonsynonymous |
| ZNF292 | | Frameshift/stop gain/splice site, Nonsynonymous |
