## Supplementary Table S3 for "Distinct characteristics of lymphoid and myeloid clonal hematopoiesis in World Trade Center first responders"

**Supplementary Table S3a: M-CHIP Mutations identified in WTC samples**

| | Chr | GrCh38 Pos | Id | Ref | Alt | Gene | Type_Var | VAF |
|---|---|---|---|---|---|---|---|---|
| 1 | chr1 | 1815790 | rs141326438 | T | C | GNB1 | nonsynonymous SNV | 0.03 |
| 2 | chr1 | 1815790 | rs141326438 | T | C | GNB1 | nonsynonymous SNV | 0.153 |
| 3 | chr1 | 114716126 | rs121913237 | C | T | NRAS | nonsynonymous SNV | 0.082 |
| 4 | chr2 | 25234373 | rs147001633 | C | T | DNMT3A | nonsynonymous SNV | 0.078 |
| 5 | chr2 | 25234374 | rs377577594 | G | A | DNMT3A | nonsynonymous SNV | 0.256 |
| 6 | chr2 | 25235724 | rs376830288 | C | T | DNMT3A | stopgain | 0.041 |
| 7 | chr2 | 25235792 | chr2_25235792_T_C | T | C | DNMT3A | nonsynonymous SNV | 0.278 |
| 8 | chr2 | 25240312 | chr2_25240312_C_T | C | T | DNMT3A | nonsynonymous SNV | 0.049 |
| 9 | chr2 | 25240313 | chr2_25240313_G_A | G | A | DNMT3A | stopgain | 0.04 |
| 10 | chr2 | 25240360 | chr2_25240360_A_G | A | G | DNMT3A | nonsynonymous SNV | 0.032 |
| 11 | chr2 | 25240371 | chr2_25240371_G_C | G | C | DNMT3A | nonsynonymous SNV | 0.026 |
| 12 | chr2 | 25240420 | rs147828672 | T | C | DNMT3A | nonsynonymous SNV | 0.038 |
| 13 | chr2 | 25240667 | chr2_25240667_C_T | C | T | DNMT3A | nonsynonymous SNV | 0.024 |
| 14 | chr2 | 25240693 | chr2_25240693_C_T | C | T | DNMT3A | nonsynonymous SNV | 0.155 |
| 15 | chr2 | 25241561 | chr2_25241561_C_A | C | A | DNMT3A | splicing | 0.038 |
| 16 | chr2 | 25241601 | chr2_25241600_TG_T | G | - | DNMT3A | frameshift deletion | 0.236 |
| 17 | chr2 | 25241664 | chr2_25241664_G_C | G | C | DNMT3A | stopgain | 0.044 |
| 18 | chr2 | 25243921 | chr2_25243921_G_A | G | A | DNMT3A | nonsynonymous SNV | 0.191 |
| 19 | chr2 | 25244203 | chr2_25244203_C_T | C | T | DNMT3A | stopgain | 0.035 |
| 20 | chr2 | 25246186 | chr2_25246186_T_C | T | C | DNMT3A | nonsynonymous SNV | 0.103 |
| 21 | chr2 | 25247063 | chr2_25247063_G_C | G | C | DNMT3A | stopgain | 0.13 |
| 22 | chr2 | 25247127 | chr2_25247127_G_T | G | T | DNMT3A | stopgain | 0.053 |
| 23 | chr2 | 25247666 | chr2_25247666_C_T | C | T | DNMT3A | stopgain | 0.031 |
| 24 | chr2 | 25247751 | chr2_25247751_T_C | T | C | DNMT3A | splicing | 0.04 |
| 25 | chr2 | 25274945 | chr2_25274944_CCT_C | CT | - | DNMT3A | frameshift deletion | 0.039 |
| 26 | chr2 | 197402110 | chr2_197402110_T_C | T | C | SF3B1 | nonsynonymous SNV | 0.051 |
| 27 | chr2 | 197402110 | chr2_197402110_T_C | T | C | SF3B1 | nonsynonymous SNV | 0.112 |
| 28 | chr2 | 197402636 | chr2_197402636_T_C | T | C | SF3B1 | nonsynonymous SNV | 0.332 |
| 29 | chr4 | 105234317 | chr4_105234316_TC_T | C | - | TET2 | frameshift deletion | 0.106 |
| 30 | chr4 | 105235101 | chr4_105235100_GTTCACTAA | TTCACTAAG | - | TET2 | frameshift deletion | 0.022 |
| 31 | chr4 | 105235500 | chr4_105235499_TG_T | G | - | TET2 | frameshift deletion | 0.142 |
| 32 | chr4 | 105236201 | chr4_105236200_AAG_A | AG | - | TET2 | frameshift deletion | 0.064 |
| 33 | chr4 | 105236592 | chr4_105236592_C_T | C | T | TET2 | stopgain | 0.033 |
| 34 | chr4 | 105242916 | chr4_105242916_A_G | A | G | TET2 | nonsynonymous SNV | 0.409 |
| 35 | chr4 | 105242921 | chr4_105242920_CTAAGTGG | TAAGTGGG | - | TET2 | frameshift deletion | 0.034 |
| 36 | chr4 | 105243621 | chr4_105243621_C_T | C | T | TET2 | stopgain | 0.033 |
| 37 | chr4 | 105243760 | chr4_105243760_G_A | G | A | TET2 | nonsynonymous SNV | 0.042 |
| 38 | chr4 | 105259675 | chr4_105259675_T_G | T | G | TET2 | nonsynonymous SNV | 0.072 |
| 39 | chr4 | 105261769 | chr4_105261769_T_C | T | C | TET2 | nonsynonymous SNV | 0.289 |
| 40 | chr4 | 105269661 | chr4_105269661_C_T | C | T | TET2 | nonsynonymous SNV | 0.431 |
| 41 | chr4 | 105272673 | chr4_105272673_G_A | G | A | TET2 | nonsynonymous SNV | 0.021 |
| 42 | chr4 | 105275056 | rs370735654 | C | T | TET2 | stopgain | 0.046 |
| 43 | chr4 | 105275793 | chr4_105275792_AC_A | C | - | TET2 | frameshift deletion | 0.323 |
| 44 | chr4 | 105275794 | chr4_105275794_A_ATG | - | TG | TET2 | frameshift insertion | 0.323 |
| 45 | chr4 | 105276113 | chr4_105276113_A_G | A | G | TET2 | nonsynonymous SNV | 0.032 |
| 46 | chr6 | 79026017 | chr6_79026017_G_A | G | A | PHIP | stopgain | 0.036 |
| 47 | chr7 | 102196694 | chr7_102196693_GC_G | C | - | CUX1 | frameshift deletion | 0.034 |
| 48 | chr9 | 5073770 | rs77375493 | G | T | JAK2 | nonsynonymous SNV | 0.24 |
| 49 | chr11 | 32389180 | chr11_32389180_C_T | C | T | WT1 | splicing | 0.032 |
| 50 | chr17 | 7676257 | chr17_7676256_TGGGAC_T | GGGAC | - | TP53 | frameshift deletion | 0.027 |
| 51 | chr17 | 60656823 | chr17_60656823_T_A | T | A | PPM1D | stopgain | 0.031 |
| 52 | chr17 | 60663076 | chr17_60663075_GA_G | A | - | PPM1D | frameshift deletion | 0.043 |
| 53 | chr17 | 60663078 | chr17_60663077_AT_A | T | - | PPM1D | frameshift deletion | 0.085 |
| 54 | chr17 | 60663114 | chr17_60663113_AT_A | T | - | PPM1D | frameshift deletion | 0.114 |
| 55 | chr17 | 60663144 | chr17_60663143_AT_A | T | - | PPM1D | frameshift deletion | 0.029 |
| 56 | chr17 | 60663157 | chr17_60663157_G_T | G | T | PPM1D | stopgain | 0.025 |
| 57 | chr17 | 60663165 | chr17_60663165_T_TG | - | G | PPM1D | frameshift insertion | 0.055 |
| 58 | chr17 | 60663283 | chr17_60663282_AAC_A | AC | - | PPM1D | frameshift deletion | 0.026 |
| 59 | chr17 | 60663388 | chr17_60663388_C_T | C | T | PPM1D | stopgain | 0.038 |

| | | | | | | | | |
|---|---|---|---|---|---|---|---|---|
| 60 | chr17 | 60663388 | chr17_60663388_C_T | C | T | PPM1D | stopgain | 0.03 |
| 61 | chr17 | 60663448 | chr17_60663448_C_T | C | T | PPM1D | stopgain | 0.033 |
| 62 | chr17 | 76736877 | chr17_76736877_G_T | G | T | SRSF2 | nonsynonymous SNV | 0.058 |
| 63 | chr17 | 76736877 | chr17_76736877_G_A | G | A | SRSF2 | nonsynonymous SNV | 0.022 |
| 64 | chr17 | 76736877 | chr17_76736877_G_T | G | T | SRSF2 | nonsynonymous SNV | 0.042 |
| 65 | chr20 | 32433480 | chr20_32433480_C_T | C | T | ASXL1 | stopgain | 0.041 |
| 66 | chr20 | 32433807 | chr20_32433807_G_T | G | T | ASXL1 | stopgain | 0.395 |
| 67 | chr20 | 32434485 | rs371369583 | C | G | ASXL1 | stopgain | 0.047 |
| 68 | chr20 | 32434798 | chr20_32434798_C_CTCAG | - | TCAG | ASXL1 | frameshift insertion | 0.036 |
| 69 | chr20 | 32435100 | chr20_32435100_G_A | G | A | ASXL1 | stopgain | 0.028 |
| 70 | chr20 | 32435356 | chr20_32435356_C_T | C | T | ASXL1 | stopgain | 0.042 |
| 71 | chr20 | 32435834 | chr20_32435833_GC_G | C | - | ASXL1 | frameshift deletion | 0.398 |

**Supplementary Table S3b: L-CHIP Mutations identified in WTC samples**

| | Chr | GrCh38 Pos | Id | Ref | Alt | Gene | Type_Var | VAF |
|---|---|---|---|---|---|---|---|---|
| 1 | chr1 | 11253909 | chr1_11253909_C_T | C | T | MTOR | nonsynonymous SNV | 0.03 |
| 2 | chr1 | 26779440 | chr1_26779439_TG_T | G | - | ARID1A | frameshift deletion | 0.028 |
| 3 | chr1 | 144437970 | chr1_144437970_G_A | G | A | NBPF15 | stopgain | 0.075 |
| 4 | chr1 | 150579037 | chr1_150579037_G_T | G | T | MCL1 | nonsynonymous SNV | 0.085 |
| 5 | chr2 | 159386588 | chr2_159386588_C_T | C | T | BAZ2B | nonsynonymous SNV | 0.066 |
| 6 | chr2 | 237350200 | chr2_237350200_C_T | C | T | COL6A3 | nonsynonymous SNV | 0.143 |
| 7 | chr3 | 7740378 | chr3_7740378_A_G | A | G | GRM7 | nonsynonymous SNV | 0.072 |
| 8 | chr3 | 9475103 | chr3_9475103_T_C | T | C | SETD5 | nonsynonymous SNV | 0.044 |
| 9 | chr4 | 152352543 | chr4_152352543_C_T | C | T | FBXW7 | nonsynonymous SNV | 0.03 |
| 10 | chr4 | 186619480 | rs201741692 | G | A | FAT1 | nonsynonymous SNV | 0.066 |
| 11 | chr5 | 151544477 | chr5_151544477_G_A | G | A | FAT2 | nonsynonymous SNV | 0.026 |
| 12 | chr5 | 151545563 | chr5_151545563_G_A | G | A | FAT2 | nonsynonymous SNV | 0.034 |
| 13 | chr5 | 151566767 | rs142276415 | A | G | FAT2 | nonsynonymous SNV | 0.026 |
| 14 | chr5 | 157087213 | chr5_157087213_A_T | A | T | HAVCR2 | stopgain | 0.043 |
| 15 | chr5 | 157222947 | rs146738280 | C | T | ITK | nonsynonymous SNV | 0.103 |
| 16 | chr5 | 158714167 | chr5_158714167_T_C | T | C | EBF1 | nonsynonymous SNV | 0.032 |
| 17 | chr6 | 33670455 | rs200526442 | C | T | ITPR3 | nonsynonymous SNV | 0.027 |
| 18 | chr6 | 35459439 | chr6_35459439_C_T | C | T | FANCE | stopgain | 0.076 |
| 19 | chr6 | 73518440 | chr6_73518440_C_G | C | G | EEF1A1 | nonsynonymous SNV | 0.101 |
| 20 | chr6 | 73518440 | chr6_73518440_C_G | C | G | EEF1A1 | nonsynonymous SNV | 0.071 |
| 21 | chr6 | 73518440 | chr6_73518440_C_G | C | G | EEF1A1 | nonsynonymous SNV | 0.18 |
| 22 | chr6 | 73518440 | chr6_73518440_C_G | C | G | EEF1A1 | nonsynonymous SNV | 0.09 |
| 23 | chr6 | 73518440 | chr6_73518440_C_G | C | G | EEF1A1 | nonsynonymous SNV | 0.045 |
| 24 | chr6 | 73518506 | chr6_73518506_C_T | C | T | EEF1A1 | nonsynonymous SNV | 0.034 |
| 25 | chr6 | 73518506 | chr6_73518506_C_T | C | T | EEF1A1 | nonsynonymous SNV | 0.08 |
| 26 | chr6 | 73518506 | chr6_73518506_C_T | C | T | EEF1A1 | nonsynonymous SNV | 0.056 |
| 27 | chr6 | 73518506 | chr6_73518506_C_T | C | T | EEF1A1 | nonsynonymous SNV | 0.079 |
| 28 | chr6 | 73518506 | chr6_73518506_C_T | C | T | EEF1A1 | nonsynonymous SNV | 0.082 |
| 29 | chr6 | 73518506 | chr6_73518506_C_T | C | T | EEF1A1 | nonsynonymous SNV | 0.09 |
| 30 | chr6 | 73518506 | chr6_73518506_C_T | C | T | EEF1A1 | nonsynonymous SNV | 0.087 |
| 31 | chr6 | 73518506 | chr6_73518506_C_T | C | T | EEF1A1 | nonsynonymous SNV | 0.068 |
| 32 | chr6 | 73518506 | chr6_73518506_C_T | C | T | EEF1A1 | nonsynonymous SNV | 0.082 |
| 33 | chr6 | 73518506 | chr6_73518506_C_T | C | T | EEF1A1 | nonsynonymous SNV | 0.069 |
| 34 | chr6 | 73518506 | chr6_73518506_C_T | C | T | EEF1A1 | nonsynonymous SNV | 0.055 |
| 35 | chr6 | 73518506 | chr6_73518506_C_T | C | T | EEF1A1 | nonsynonymous SNV | 0.086 |
| 36 | chr6 | 73518506 | chr6_73518506_C_T | C | T | EEF1A1 | nonsynonymous SNV | 0.058 |
| 37 | chr6 | 75256280 | chr6_75256280_T_G | T | G | TMEM30A | nonsynonymous SNV | 0.03 |
| 38 | chr6 | 75284514 | chr6_75284514_T_C | T | C | TMEM30A | nonsynonymous SNV | 0.046 |
| 39 | chr6 | 134174532 | chr6_134174532_T_C | T | C | SGK1 | nonsynonymous SNV | 0.052 |
| 40 | chr8 | 10623053 | chr8_10623053_C_T | C | T | RP1L1 | nonsynonymous SNV | 0.136 |
| 41 | chr8 | 102287474 | chr8_102287474_C_T | C | T | UBR5 | nonsynonymous SNV | 0.035 |
| 42 | chr8 | 102311477 | chr8_102311477_C_A | C | A | UBR5 | nonsynonymous SNV | 0.029 |
| 43 | chr9 | 95469004 | chr9_95469004_G_A | G | A | PTCH1 | nonsynonymous SNV | 0.03 |
| 44 | chr9 | 130873007 | chr9_130873007_A_G | A | G | ABL1 | nonsynonymous SNV | 0.036 |
| 45 | chr11 | 102325266 | chr11_102325266_C_T | C | T | BIRC3 | stopgain | 0.491 |
| 46 | chr11 | 108310209 | chr11_108310209_T_A | T | A | ATM | nonsynonymous SNV | 0.068 |
| 47 | chr11 | 108327754 | chr11_108327753_GA_G | A | - | ATM | frameshift deletion | 0.5 |
| 48 | chr11 | 108330292 | chr11_108330292_C_G | C | G | ATM | nonsynonymous SNV | 0.039 |
| 49 | chr12 | 31091731 | rs201612562 | C | T | DDX11 | nonsynonymous SNV | 0.039 |
| 50 | chr12 | 31091731 | rs201612562 | C | T | DDX11 | nonsynonymous SNV | 0.061 |
| 51 | chr12 | 31091731 | rs201612562 | C | T | DDX11 | nonsynonymous SNV | 0.087 |
| 52 | chr12 | 31091731 | rs201612562 | C | T | DDX11 | nonsynonymous SNV | 0.043 |
| 53 | chr12 | 31091731 | rs201612562 | C | T | DDX11 | nonsynonymous SNV | 0.044 |
| 54 | chr12 | 31091731 | rs201612562 | C | T | DDX11 | nonsynonymous SNV | 0.087 |
| 55 | chr12 | 31091731 | rs201612562 | C | T | DDX11 | nonsynonymous SNV | 0.088 |
| 56 | chr12 | 31091731 | rs201612562 | C | T | DDX11 | nonsynonymous SNV | 0.101 |
| 57 | chr12 | 31091731 | rs201612562 | C | T | DDX11 | nonsynonymous SNV | 0.06 |
| 58 | chr12 | 31091731 | rs201612562 | C | T | DDX11 | nonsynonymous SNV | 0.054 |
| 59 | chr12 | 31091731 | rs201612562 | C | T | DDX11 | nonsynonymous SNV | 0.07 |

| | | | | | | | | |
|---|---|---|---|---|---|---|---|---|
| 60 | chr12 | 31096347 | chr12_31096347_C_T | C | T | DDX11 | stopgain | 0.106 |
| 61 | chr12 | 31100666 | chr12_31100666_G_C | G | C | DDX11 | nonsynonymous SNV | 0.049 |
| 62 | chr12 | 49040415 | chr12_49040415_C_T | C | T | KMT2D | nonsynonymous SNV | 0.035 |
| 63 | chr12 | 49041225 | chr12_49041225_G_A | G | A | KMT2D | nonsynonymous SNV | 0.026 |
| 64 | chr12 | 49044253 | chr12_49044253_T_C | T | C | KMT2D | nonsynonymous SNV | 0.028 |
| 65 | chr12 | 49044253 | chr12_49044253_T_C | T | C | KMT2D | nonsynonymous SNV | 0.028 |
| 66 | chr12 | 49051025 | chr12_49051025_A_AG | - | G | KMT2D | frameshift insertion | 0.377 |
| 67 | chr12 | 56599190 | chr12_56599190_A_G | A | G | BAZ2A | nonsynonymous SNV | 0.029 |
| 68 | chr12 | 124340312 | chr12_124340312_C_T | C | T | NCOR2 | nonsynonymous SNV | 0.026 |
| 69 | chr12 | 124372307 | chr12_124372307_T_C | T | C | NCOR2 | nonsynonymous SNV | 0.032 |
| 70 | chr13 | 48377030 | chr13_48377030_C_A | C | A | RB1 | stopgain | 0.766 |
| 71 | chr14 | 21431453 | chr14_21431453_C_T | C | T | CHD8 | nonsynonymous SNV | 0.027 |
| 72 | chr14 | 32732491 | chr14_32732491_A_G | A | G | AKAP6 | nonsynonymous SNV | 0.032 |
| 73 | chr15 | 41711188 | chr15_41711188_C_T | C | T | MGA | stopgain | 0.028 |
| 74 | chr15 | 75400915 | chr15_75400915_A_G | A | G | SIN3A | nonsynonymous SNV | 0.039 |
| 75 | chr15 | 92927325 | chr15_92927325_G_T | G | T | CHD2 | stopgain | 0.104 |
| 76 | chr15 | 92992953 | chr15_92992953_G_A | G | A | CHD2 | nonsynonymous SNV | 0.04 |
| 77 | chr16 | 81956840 | chr16_81956840_A_G | A | G | PLCG2 | nonsynonymous SNV | 0.037 |
| 78 | chr17 | 42322464 | chr17_42322464_T_A | T | A | STAT3 | nonsynonymous SNV | 0.037 |
| 79 | chr17 | 63929438 | chr17_63929438_T_C | T | C | CD79B | nonsynonymous SNV | 0.503 |
| 80 | chr17 | 75393616 | chr17_75393616_C_T | C | T | GRB2 | nonsynonymous SNV | 0.027 |
| 81 | chr19 | 1434845 | chr19_1434845_C_T | C | T | DAZAP1 | nonsynonymous SNV | 0.043 |
| 82 | chr19 | 42092192 | chr19_42092192_G_A | G | A | POU2F2 | nonsynonymous SNV | 0.06 |
| 83 | chr19 | 42095341 | chr19_42095341_C_G | C | G | POU2F2 | nonsynonymous SNV | 0.059 |
| 84 | chrX | 32380604 | chrX_32380604_G_C | G | C | DMD | nonsynonymous SNV | 0.19 |
| 85 | chrX | 41341579 | chrX_41341579_A_G | A | G | DDX3X | nonsynonymous SNV | 0.097 |

**Supplementary Table S3c: Distribution of M-CHIP mutations by genes**

| Gene | # Mutations | # Individuals |
|---|---|---|
| DNMT3A | 22 | 22 |
| TET2 | 17 | 15 |
| PPM1D | 11 | 11 |
| ASXL1 | 7 | 7 |
| SF3B1 | 3 | 3 |
| SRSF2 | 3 | 3 |
| GNB1 | 2 | 2 |
| CUX1 | 1 | 1 |
| JAK2 | 1 | 1 |
| NRAS | 1 | 1 |
| PHIP | 1 | 1 |
| TP53 | 1 | 1 |
| WT1 | 1 | 1 |

**Supplementary Table S3d: Distribution of L-CHIP mutations by genes**

| Gene | # Mutations | # Individuals |
|---|---|---|
| EEF1A1 | 18 | 18 |
| DDX11 | 13 | 13 |
| KMT2D | 5 | 5 |
| ATM | 3 | 3 |
| FAT2 | 3 | 3 |
| CHD2 | 2 | 2 |
| NCOR2 | 2 | 1 |
| POU2F2 | 2 | 2 |
| TMEM30A | 2 | 2 |
| UBR5 | 2 | 2 |
| ABL1 | 1 | 1 |
| AKAP6 | 1 | 1 |
| ARID1A | 1 | 1 |
| BAZ2A | 1 | 1 |
| BAZ2B | 1 | 1 |
| BIRC3 | 1 | 1 |
| CD79B | 1 | 1 |
| CHD8 | 1 | 1 |
| COL6A3 | 1 | 1 |
| DAZAP1 | 1 | 1 |
| DDX3X | 1 | 1 |
| DMD | 1 | 1 |
| EBF1 | 1 | 1 |
| FANCE | 1 | 1 |
| FAT1 | 1 | 1 |
| FBXW7 | 1 | 1 |
| GRB2 | 1 | 1 |
| GRM7 | 1 | 1 |
| HAVCR2 | 1 | 1 |
| ITK | 1 | 1 |
| ITPR3 | 1 | 1 |
| MCL1 | 1 | 1 |
| MGA | 1 | 1 |
| MTOR | 1 | 1 |
| NBPF15 | 1 | 1 |
| PLCG2 | 1 | 1 |
| PTCH1 | 1 | 1 |
| RB1 | 1 | 1 |
| RP1L1 | 1 | 1 |
| SETD5 | 1 | 1 |
| SGK1 | 1 | 1 |
| SIN3A | 1 | 1 |
| STAT3 | 1 | 1 |

**Supplementary Table S3e: M-CHIP Mutations identified in downsampled WTC samples**

| | Chr | GrCh38 Pos | Id | Ref | Alt | Gene | Type_Var | VAF |
|---|---|---|---|---|---|---|---|---|
| 1 | chr1 | 1815790 | rs141326438 | T | C | GNB1 | nonsynonymous SNV | 0.2 |
| 2 | chr2 | 25234374 | rs377577594 | G | A | DNMT3A | nonsynonymous SNV | 0.205 |
| 3 | chr2 | 25235724 | rs376830288 | C | T | DNMT3A | stopgain | 0.052 |
| 4 | chr2 | 25235792 | chr2_25235792_T_C | T | C | DNMT3A | nonsynonymous SNV | 0.316 |
| 5 | chr2 | 25240312 | chr2_25240312_C_T | C | T | DNMT3A | nonsynonymous SNV | 0.051 |
| 6 | chr2 | 25240313 | chr2_25240313_G_A | G | A | DNMT3A | stopgain | 0.083 |
| 7 | chr2 | 25240693 | chr2_25240693_C_T | C | T | DNMT3A | nonsynonymous SNV | 0.111 |
| 8 | chr2 | 25241601 | chr2_25241600_TG_T | G | - | DNMT3A | frameshift deletion | 0.225 |
| 9 | chr2 | 25243921 | chr2_25243921_G_A | G | A | DNMT3A | nonsynonymous SNV | 0.155 |
| 10 | chr2 | 25244203 | chr2_25244203_C_T | C | T | DNMT3A | stopgain | 0.063 |
| 11 | chr2 | 25246186 | chr2_25246186_T_C | T | C | DNMT3A | nonsynonymous SNV | 0.1 |
| 12 | chr2 | 25247063 | chr2_25247063_G_C | G | C | DNMT3A | stopgain | 0.226 |
| 13 | chr2 | 25247076 | chr2_25247076_C_T | C | T | DNMT3A | nonsynonymous SNV | 0.079 |
| 14 | chr2 | 197402110 | chr2_197402110_T_C | T | C | SF3B1 | nonsynonymous SNV | 0.148 |
| 15 | chr2 | 197402110 | chr2_197402110_T_C | T | C | SF3B1 | nonsynonymous SNV | 0.275 |
| 16 | chr4 | 105234317 | chr4_105234316_TC_T | C | - | TET2 | frameshift deletion | 0.071 |
| 17 | chr4 | 105235500 | chr4_105235499_TG_T | G | - | TET2 | frameshift deletion | 0.169 |
| 18 | chr4 | 105236201 | chr4_105236200_AAG_A | AG | - | TET2 | frameshift deletion | 0.091 |
| 19 | chr4 | 105242916 | chr4_105242916_A_G | A | G | TET2 | nonsynonymous SNV | 0.439 |
| 20 | chr4 | 105259675 | chr4_105259675_T_G | T | G | TET2 | nonsynonymous SNV | 0.064 |
| 21 | chr4 | 105261769 | chr4_105261769_T_C | T | C | TET2 | nonsynonymous SNV | 0.289 |
| 22 | chr4 | 105269661 | chr4_105269661_C_T | C | T | TET2 | nonsynonymous SNV | 0.424 |
| 23 | chr4 | 105272909 | chr4_105272909_C_T | C | T | TET2 | stopgain | 0.063 |
| 24 | chr4 | 105275056 | rs370735654 | C | T | TET2 | stopgain | 0.075 |
| 25 | chr4 | 105275793 | chr4_105275792_AC_A | C | - | TET2 | frameshift deletion | 0.312 |
| 26 | chr4 | 105275794 | chr4_105275794_A_ATG | - | TG | TET2 | frameshift insertion | 0.312 |
| 27 | chr16 | 3739726 | chr16_3739726_T_C | T | C | CREBBP | splicing | 0.075 |
| 28 | chr17 | 60663114 | chr17_60663113_AT_A | T | - | PPM1D | frameshift deletion | 0.111 |
| 29 | chr17 | 60663165 | chr17_60663165_T_TG | - | G | PPM1D | frameshift insertion | 0.059 |
| 30 | chr17 | 60663388 | chr17_60663388_C_T | C | T | PPM1D | stopgain | 0.058 |
| 31 | chr17 | 76736877 | chr17_76736877_G_T | G | T | SRSF2 | nonsynonymous SNV | 0.075 |
| 32 | chr20 | 32433807 | chr20_32433807_G_T | G | T | ASXL1 | stopgain | 0.388 |
| 33 | chr20 | 32435100 | chr20_32435100_G_A | G | A | ASXL1 | stopgain | 0.049 |

**Supplementary Table S3f: L-CHIP Mutations identified in downsampled WTC samples**

| | Chr | GrCh38 Pos | Id | Ref | Alt | Gene | Type_Var | VAF |
|---|---|---|---|---|---|---|---|---|
| 1 | chr1 | 88983034 | rs202218737 | C | T | RBMXL1 | nonsynonymous SNV | 0.037 |
| 2 | chr1 | 144437095 | chr1_144437095_A_G | A | G | NBPF15 | nonsynonymous SNV | 0.039 |
| 3 | chr1 | 150579037 | chr1_150579037_G_T | G | T | MCL1 | nonsynonymous SNV | 0.079 |
| 4 | chr2 | 159386588 | chr2_159386588_C_T | C | T | BAZ2B | nonsynonymous SNV | 0.071 |
| 5 | chr2 | 237350200 | chr2_237350200_C_T | C | T | COL6A3 | nonsynonymous SNV | 0.127 |
| 6 | chr2 | 237378938 | rs370719148 | G | A | COL6A3 | nonsynonymous SNV | 0.049 |
| 7 | chr3 | 7740378 | chr3_7740378_A_G | A | G | GRM7 | nonsynonymous SNV | 0.098 |
| 8 | chr5 | 151566866 | chr5_151566866_A_G | A | G | FAT2 | nonsynonymous SNV | 0.04 |
| 9 | chr6 | 73518440 | chr6_73518440_C_G | C | G | EEF1A1 | nonsynonymous SNV | 0.093 |
| 10 | chr6 | 73518440 | chr6_73518440_C_G | C | G | EEF1A1 | nonsynonymous SNV | 0.113 |
| 11 | chr6 | 73518506 | chr6_73518506_C_T | C | T | EEF1A1 | nonsynonymous SNV | 0.14 |
| 12 | chr6 | 73518506 | chr6_73518506_C_T | C | T | EEF1A1 | nonsynonymous SNV | 0.06 |
| 13 | chr6 | 73518506 | chr6_73518506_C_T | C | T | EEF1A1 | nonsynonymous SNV | 0.072 |
| 14 | chr6 | 73518506 | chr6_73518506_C_T | C | T | EEF1A1 | nonsynonymous SNV | 0.106 |
| 15 | chr6 | 73518506 | chr6_73518506_C_T | C | T | EEF1A1 | nonsynonymous SNV | 0.091 |
| 16 | chr6 | 73518506 | chr6_73518506_C_T | C | T | EEF1A1 | nonsynonymous SNV | 0.085 |
| 17 | chr6 | 73518506 | chr6_73518506_C_T | C | T | EEF1A1 | nonsynonymous SNV | 0.096 |
| 18 | chr6 | 75284514 | chr6_75284514_T_C | T | C | TMEM30A | nonsynonymous SNV | 0.118 |
| 19 | chr6 | 134174532 | chr6_134174532_T_C | T | C | SGK1 | nonsynonymous SNV | 0.071 |
| 20 | chr6 | 152329857 | chr6_152329857_G_A | G | A | SYNE1 | nonsynonymous SNV | 0.039 |
| 21 | chr8 | 10623053 | chr8_10623053_C_T | C | T | RP1L1 | nonsynonymous SNV | 0.147 |
| 22 | chr8 | 13099855 | chr8_13099855_A_G | A | G | DLC1 | nonsynonymous SNV | 0.04 |
| 23 | chr11 | 102325266 | chr11_102325266_C_T | C | T | BIRC3 | stopgain | 0.468 |
| 24 | chr12 | 31091731 | rs201612562 | C | T | DDX11 | nonsynonymous SNV | 0.075 |
| 25 | chr12 | 31091731 | rs201612562 | C | T | DDX11 | nonsynonymous SNV | 0.081 |
| 26 | chr12 | 31091731 | rs201612562 | C | T | DDX11 | nonsynonymous SNV | 0.128 |
| 27 | chr12 | 31091731 | rs201612562 | C | T | DDX11 | nonsynonymous SNV | 0.128 |
| 28 | chr12 | 31091731 | rs201612562 | C | T | DDX11 | nonsynonymous SNV | 0.081 |
| 29 | chr12 | 31091731 | rs201612562 | C | T | DDX11 | nonsynonymous SNV | 0.086 |
| 30 | chr12 | 31091791 | chr12_31091791_C_G | C | G | DDX11 | nonsynonymous SNV | 0.119 |
| 31 | chr12 | 31096347 | chr12_31096347_C_T | C | T | DDX11 | stopgain | 0.136 |
| 32 | chr12 | 49041225 | chr12_49041225_G_A | G | A | KMT2D | nonsynonymous SNV | 0.056 |
| 33 | chr13 | 48377030 | chr13_48377030_C_A | C | A | RB1 | stopgain | 0.808 |
| 34 | chr15 | 92927325 | chr15_92927325_G_T | G | T | CHD2 | stopgain | 0.127 |
| 35 | chr17 | 42322464 | chr17_42322464_T_A | T | A | STAT3 | nonsynonymous SNV | 0.092 |
| 36 | chr17 | 63929438 | chr17_63929438_T_C | T | C | CD79B | nonsynonymous SNV | 0.588 |
| 37 | chr19 | 19145922 | chr19_19145922_C_G | C | G | MEF2B | nonsynonymous SNV | 0.071 |
| 38 | chr19 | 42095341 | chr19_42095341_C_G | C | G | POU2F2 | nonsynonymous SNV | 0.089 |
| 39 | chrX | 41341579 | chrX_41341579_A_G | A | G | DDX3X | nonsynonymous SNV | 0.151 |

**Supplementary Table S3g: M-CHIP Mutations identified in controls**

| | Chr | GrCh38 Pos | Id | Ref | Alt | Gene | Type_Var | VAF |
|---|---|---|---|---|---|---|---|---|
| 1 | chr2 | 25234312 | chr2_25234311_CG_C | G | - | DNMT3A | frameshift deletion | 0.116 |
| 2 | chr2 | 25237006 | chr2_25237006_C_T | C | T | DNMT3A | splicing | 0.3 |
| 3 | chr2 | 25244546 | chr2_25244545_GCAGT_G | CAGT | - | DNMT3A | frameshift deletion | 0.127 |
| 4 | chr2 | 25247719 | chr2_25247719_C_T | C | T | DNMT3A | nonsynonymous SNV | 0.076 |
| 5 | chr2 | 197401882 | chr2_197401882_C_G | C | G | SF3B1 | nonsynonymous SNV | 0.1 |
| 6 | chr4 | 105272631 | chr4_105272631_T_C | T | C | TET2 | nonsynonymous SNV | 0.477 |
| 7 | chr11 | 64778327 | chr11_64778326_CG_C | G | - | SF1 | frameshift deletion | 0.068 |
| 8 | chr16 | 3728047 | chr16_3728047_G_A | G | A | CREBBP | stopgain | 0.043 |
| 9 | chr20 | 32433447 | rs375215583 | C | T | ASXL1 | stopgain | 0.173 |

**Supplementary Table S3h: L-CHIP Mutations identified in controls**

| | Chr | GrCh38 Pos | Id | Ref | Alt | Gene | Type_Var | VAF |
|---|---|---|---|---|---|---|---|---|
| 1 | chr4 | 25847507 | rs370424215 | G | A | SEL1L3 | nonsynonymous SNV | 0.082 |
| 2 | chr8 | 10612540 | chr8_10612540_G_A | G | A | RP1L1 | nonsynonymous SNV | 0.043 |
| 3 | chr11 | 108247072 | rs202160435 | G | A | ATM | nonsynonymous SNV | 0.365 |
| 4 | chr12 | 31091731 | rs201612562 | C | T | DDX11 | nonsynonymous SNV | 0.145 |
| 5 | chr12 | 49042846 | chr12_49042846_G_A | G | A | KMT2D | stopgain | 0.064 |
| 6 | chr19 | 41894301 | chr19_41894300_GA_G | A | - | ARHGEF1 | frameshift deletion | 0.096 |
