## Supplementary Table S4 for "Distinct characteristics of lymphoid and myeloid clonal hematopoiesis in World Trade Center first responders"

**Supplementary Table S4a: CHIP-phenotype associations**

| Categorical Variable, N (%) | Characteristic | Negative (N=227) | CHIP positive (N=118) | (marginal) | | FDR | OR | 95% CI | Multivariate GLM |
|---|---|---|---|---|---|---|---|---|---|
| Gender | F | 18 (0.0793) | 8 (0.0678) | 0.831 | | 0.913 | | | |
| | M | 209 (0.921) | 110 (0.932) | | | | 1.18 | (0.514,2.97) | |
| Smoking status | Never Smoked | 125 (0.551) | 51 (0.432) | 0.0647 | | 0.454 | | | |
| | Previous Smoker | 91 (0.401) | 63 (0.534) | | | | 1.7 | (1.08,2.69) | |
| | Current Smoker | 11 (0.0485) | 4 (0.0339) | | | | 0.891 | (0.238,2.74) | |
| Exposure | <= Low | 34 (0.15) | 22 (0.186) | 0.462 | | 0.844 | | | |
| | Intermediate | 139 (0.612) | 71 (0.602) | | | | 0.789 | (0.432,1.46) | |
| | >=High | 50 (0.22) | 20 (0.169) | | | | 0.618 | (0.291,1.3) | |
| | Missing/ No record | 4 (0.0176) | 5 (0.0424) | | | | | | |
| Race | Non-White | 18 (0.0793) | 13 (0.11) | 0.427 | | 0.844 | | | |
| | White | 209 (0.921) | 105 (0.89) | | | | 0.696 | (0.33,1.5) | |
| CVD | No | 207 (0.912) | 104 (0.881) | 0.446 | | 0.844 | | | |
| | Yes | 20 (0.0881) | 14 (0.119) | | | | 1.39 | (0.664,2.85) | |
| Stroke | No | 225 (0.991) | 117 (0.992) | 1 | | 1 | | | |
| | Yes | 2 (0.00881) | 1 (0.00847) | | | | 0.962 | (0.0444,10.1) | |
| A | Het | 204 (0.899) | 104 (0.881) | 0.714 | | 0.893 | | | |
| | Hom | 23 (0.101) | 14 (0.119) | | | | 1.19 | (0.578,2.39) | |
| B | Het | 216 (0.952) | 114 (0.966) | 0.782 | | 0.905 | | | |
| | Hom | 11 (0.0485) | 4 (0.0339) | | | | 0.689 | (0.188,2.07) | |
| C | Het | 207 (0.912) | 110 (0.932) | 0.678 | | 0.893 | | | |
| | Hom | 20 (0.0881) | 8 (0.0678) | | | | 0.753 | (0.303,1.71) | |
| DRB1 | Het | 206 (0.907) | 113 (0.958) | 0.131 | | 0.551 | | | |
| | Hom | 21 (0.0925) | 5 (0.0424) | | | | 0.434 | (0.142,1.1) | |
| DQA1 | Het | 202 (0.89) | 101 (0.856) | 0.388 | | 0.844 | | | |
| | Hom | 25 (0.11) | 17 (0.144) | | | | 1.36 | (0.692,2.62) | |
| DQB1 | Het | 206 (0.907) | 99 (0.839) | 0.0756 | | 0.454 | | | |
| | Hom | 21 (0.0925) | 19 (0.161) | | | | 1.88 | (0.962,3.67) | |
| DPA1 | Het | 78 (0.344) | 46 (0.39) | 0.41 | | 0.844 | | | |
| | Hom | 149 (0.656) | 72 (0.61) | | | | 0.819 | (0.518,1.3) | |
| DPB1 | Het | 179 (0.789) | 99 (0.839) | 0.316 | | 0.844 | | | |
| | Hom | 48 (0.211) | 19 (0.161) | | | | 0.716 | (0.391,1.27) | |
| DMA | Het | 63 (0.278) | 35 (0.297) | 0.708 | | 0.893 | | | |
| | Hom | 164 (0.722) | 83 (0.703) | | | | 0.911 | (0.56,1.5) | |
| DMB | Het | 119 (0.524) | 46 (0.39) | 0.0229 | ** | 0.454 | | | 0.00888 |
| | Hom | 108 (0.476) | 72 (0.61) | | | | 1.72 | (1.1,2.72) | |
| DOA | Het | 6 (0.0264) | 3 (0.0254) | 1 | | 1 | | | |
| | Hom | 221 (0.974) | 115 (0.975) | | | | 1.04 | (0.269,5) | |
| DOB | Het | 95 (0.419) | 41 (0.347) | 0.204 | | 0.66 | | | |
| | Hom | 132 (0.581) | 77 (0.653) | | | | 1.35 | (0.855,2.16) | |
| DRA | Het | 98 (0.432) | 54 (0.458) | 0.65 | | 0.893 | | | |
| | Hom | 129 (0.568) | 64 (0.542) | | | | 0.9 | (0.576,1.41) | |

**Continuous Variable, Median (MAD)**

| | | | | | | | | | |
|---|---|---|---|---|---|---|---|---|---|
| Age | | 59.00 (4.45) | 61.00 (5.93) | 0.00216 | ** | 0.0907 | 1.42 | (1.14,1.78) | 0.000924 |
| | Missing/ No record | 0 | 0 | | | | | | |
| BMI | | 30.11 (4.95) | 29.45 (4.87) | 0.179 | | 0.66 | 0.828 | (0.654,1.04) | |
| | Missing/ No record | 0 | 0 | | | | | | |
| Total cholesterol | | 190.50 (39.29) | 191.00 (38.55) | 0.905 | | 0.95 | 1.02 | (0.797,1.3) | |
| | Missing/ No record | 35 | 21 | | | | | | |
| Triglyceride | | 119.00 (60.05) | 118.00 (65.23) | 0.746 | | 0.896 | 0.856 | (0.65,1.1) | |
| | Missing/ No record | 35 | 21 | | | | | | |
| HDL | | 47.00 (11.86) | 50.00 (13.34) | 0.0671 | | 0.454 | 1.31 | (1.03,1.68) | |
| | Missing/ No record | 35 | 21 | | | | | | |
| LDL | | 113.00 (32.62) | 105.00 (34.10) | 0.402 | | 0.844 | 0.894 | (0.696,1.14) | |
| | Missing/ No record | 40 | 21 | | | | | | |
| VLDL | | 24.00 (11.86) | 24.00 (11.86) | 0.723 | | 0.893 | 0.853 | (0.647,1.1) | |
| | Missing/ No record | 35 | 21 | | | | | | |
| PCL | | 22.00 (7.41) | 24.50 (9.64) | 0.331 | | 0.844 | 0.973 | (0.773,1.22) | |
| | Missing/ No record | 0 | 2 | | | | | | |
| MoCA | | 26.00 (2.97) | 25.00 (2.97) | 0.0572 | | 0.454 | 0.786 | (0.607,1.01) | |
| | Missing/ No record | 55 | 27 | | | | | | |
| Absolute basophil | | 0.04 (0.01) | 0.04 (0.01) | 0.563 | | 0.893 | 1.1 | (0.863,1.41) | |
| | Missing/ No record | 35 | 21 | | | | | | |
| Absolute lymphocytes | | 1.75 (0.42) | 1.67 (0.53) | 0.586 | | 0.893 | 1.08 | (0.84,1.37) | |
| | Missing/ No record | 37 | 22 | | | | | | |
| Absolute neutrophils | | 3.66 (1.10) | 3.77 (1.16) | 0.194 | | 0.66 | 1.2 | (0.94,1.53) | |
| | Missing/ No record | 35 | 21 | | | | | | |
| Segmented neutrophils | | 59.50 (8.30) | 60.05 (8.23) | 0.585 | | 0.893 | 1.09 | (0.848,1.39) | |
| | Missing/ No record | 37 | 22 | | | | | | |
| Absolute eosinophils | | 0.17 (0.10) | 0.16 (0.10) | 0.346 | | 0.844 | 0.938 | (0.721,1.2) | |
| | Missing/ No record | 35 | 22 | | | | | | |

| | | | | | | | | |
|---|---|---|---|---|---|---|---|---|
| Absolute monocytes | | 0.53 (0.13) | 0.54 (0.13) | 0.639 | | 0.893 | 1.08 | (0.843,1.39) |
| | Missing/ No record | 35 | 21 | | | | | |
| Platelet count | | 233.00 (54.86) | 225.00 (44.48) | 0.101 | | 0.489 | 0.83 | (0.642,1.06) |
| | Missing/ No record | 36 | 21 | | | | | |
| RBC | | 4.99 (0.40) | 4.91 (0.43) | 0.071 | | 0.454 | 0.888 | (0.543,1.17) |
| | Missing/ No record | 36 | 21 | | | | | |
| RDW | | 12.80 (0.74) | 12.80 (0.59) | 0.797 | | 0.905 | 0.935 | (0.715,1.19) |
| | Missing/ No record | 36 | 21 | | | | | |
| MCH | | 30.10 (1.78) | 30.30 (1.33) | 0.226 | | 0.677 | 1.2 | (0.937,1.57) |
| | Missing/ No record | 36 | 21 | | | | | |
| MCHC | | 34.10 (0.89) | 34.00 (0.59) | 0.706 | | 0.893 | 0.99 | (0.776,1.27) |
| | Missing/ No record | 36 | 21 | | | | | |
| MCV | | 88.00 (4.45) | 89.00 (4.45) | 0.105 | | 0.489 | 1.28 | (0.992,1.67) |
| | Missing/ No record | 36 | 21 | | | | | |
| WBC | | 6.20 (1.78) | 6.20 (1.78) | 0.848 | | 0.913 | 1.2 | (0.936,1.69) |
| | Missing/ No record | 36 | 21 | | | | | |
| LMR | | 3.21 (1.07) | 3.13 (1.11) | 0.489 | | 0.857 | 1.05 | (0.809,1.35) |
| | Missing/ No record | 37 | 22 | | | | | |

**Supplementary Table S4b: M-CHIP-phenotype associations**

| Categorical Variable, N (%) | Characteristic | Negative (N=227) | M-CHIP positive (N=56) | p (marginal) | | FDR | OR | 95% CI | Multivariate GLM |
|---|---|---|---|---|---|---|---|---|---|
| Gender | F | 18 (0.0793) | 3 (0.0536) | 0.776 | | 0.958 | | | |
| | M | 209 (0.921) | 53 (0.946) | | | | 1.52 | (0.492,6.67) | |
| Smoking status | Never Smoked | 125 (0.551) | 20 (0.357) | 0.0192 | ** | 0.331 | | | 0.0125 |
| | Previous Smoker | 91 (0.401) | 34 (0.607) | | | | 2.34 | (1.27,4.38) | |
| | Current Smoker | 11 (0.0485) | 2 (0.0357) | | | | 1.14 | (0.168,4.65) | |
| Exposure | <= Low | 34 (0.15) | 12 (0.214) | 0.438 | | 0.799 | | | |
| | Intermediate | 139 (0.612) | 32 (0.571) | | | | 0.652 | (0.31,1.44) | |
| | >=High | 50 (0.22) | 10 (0.179) | | | | 0.567 | (0.216,1.46) | |
| | Missing/ No record | 4 (0.0176) | 2 (0.0357) | | | | | | |
| Race | Non-White | 18 (0.0793) | 7 (0.125) | 0.295 | | 0.652 | | | |
| | White | 209 (0.921) | 49 (0.875) | | | | 0.603 | (0.247,1.62) | |
| CVD | No | 207 (0.912) | 48 (0.857) | 0.218 | | 0.61 | | | |
| | Yes | 20 (0.0881) | 8 (0.143) | | | | 1.73 | (0.68,4.03) | |
| Stroke | No | 225 (0.991) | 55 (0.982) | 0.485 | | 0.809 | | | |
| | Yes | 2 (0.00881) | 1 (0.0179) | | | | 2.05 | (0.0941,21.7) | |
| A | Het | 204 (0.899) | 51 (0.911) | 1 | | 1 | | | |
| | Hom | 23 (0.101) | 5 (0.0893) | | | | 0.87 | (0.281,2.23) | |
| B | Het | 216 (0.952) | 54 (0.964) | 1 | | 1 | | | |
| | Hom | 11 (0.0485) | 2 (0.0357) | | | | 0.727 | (0.11,2.81) | |
| C | Het | 207 (0.912) | 52 (0.929) | 1 | | 1 | | | |
| | Hom | 20 (0.0881) | 4 (0.0714) | | | | 0.796 | (0.224,2.21) | |
| DRB1 | Het | 206 (0.907) | 54 (0.964) | 0.272 | | 0.637 | | | |
| | Hom | 21 (0.0925) | 2 (0.0357) | | | | 0.363 | (0.057,1.29) | |
| DQA1 | Het | 202 (0.89) | 46 (0.821) | 0.176 | | 0.569 | | | |
| | Hom | 25 (0.11) | 10 (0.179) | | | | 1.76 | (0.759,3.82) | |
| DQB1 | Het | 206 (0.907) | 49 (0.875) | 0.458 | | 0.802 | | | |
| | Hom | 21 (0.0925) | 7 (0.125) | | | | 1.4 | (0.527,3.34) | |
| DPA1 | Het | 78 (0.344) | 26 (0.464) | 0.121 | | 0.546 | | | |
| | Hom | 149 (0.656) | 30 (0.536) | | | | 0.604 | (0.334,1.1) | |
| DPB1 | Het | 179 (0.789) | 46 (0.821) | 0.712 | | 0.935 | | | |
| | Hom | 48 (0.211) | 10 (0.179) | | | | 0.811 | (0.364,1.67) | |
| DMA | Het | 63 (0.278) | 17 (0.304) | 0.741 | | 0.943 | | | |
| | Hom | 164 (0.722) | 39 (0.696) | | | | 0.881 | (0.471,1.7) | |
| DMB | Het | 119 (0.524) | 21 (0.375) | 0.0527 | | 0.331 | | | |
| | Hom | 108 (0.476) | 35 (0.625) | | | | 1.84 | (1.02,3.39) | |
| DOA | Het | 6 (0.0264) | 1 (0.0179) | 1 | | 1 | | | |
| | Hom | 221 (0.974) | 55 (0.982) | | | | 1.49 | (0.248,28.5) | |
| DOB | Het | 95 (0.419) | 23 (0.411) | 1 | | 1 | | | |
| | Hom | 132 (0.581) | 33 (0.589) | | | | 1.03 | (0.572,1.89) | |
| DRA | Het | 98 (0.432) | 27 (0.482) | 0.549 | | 0.823 | | | |
| | Hom | 129 (0.568) | 29 (0.518) | | | | 0.816 | (0.453,1.47) | |

**Continuous Variable, Median (MAD)**

| | | | | | | | | | |
|---|---|---|---|---|---|---|---|---|---|
| Age | | 59.00 (4.45) | 62.00 (7.41) | 2.45E-05 | ** | 0.00103 | 1.81 | (1.38,2.4) | 0.0246 |
| | Missing/ No record | 0 | 0 | | | | | | |
| BMI | | 30.11 (4.95) | 28.64 (3.48) | 0.0311 | ** | 0.331 | 0.687 | (0.487,0.943) | 0.321 |
| | Missing/ No record | 0 | 0 | | | | | | |
| Total cholesterol | | 190.50 (39.29) | 194.00 (37.06) | 0.71 | | 0.935 | 0.843 | (0.566,1.24) | |
| | Missing/ No record | 35 | 14 | | | | | | |
| Triglyceride | | 119.00 (60.05) | 117.00 (65.23) | 0.52 | | 0.809 | 0.783 | (0.512,1.12) | |
| | Missing/ No record | 35 | 14 | | | | | | |
| HDL | | 47.00 (11.86) | 50.00 (13.34) | 0.132 | | 0.546 | 1.33 | (0.93,1.9) | |
| | Missing/ No record | 35 | 14 | | | | | | |
| LDL | | 113.00 (32.62) | 103.00 (34.10) | 0.365 | | 0.704 | 0.834 | (0.586,1.17) | |
| | Missing/ No record | 40 | 14 | | | | | | |
| VLDL | | 24.00 (11.86) | 23.50 (12.60) | 0.502 | | 0.809 | 0.782 | (0.51,1.11) | |
| | Missing/ No record | 35 | 14 | | | | | | |
| PCL | | 22.00 (7.41) | 22.00 (7.41) | 0.573 | | 0.83 | 0.805 | (0.568,1.09) | |
| | Missing/ No record | 0 | 0 | | | | | | |
| MoCA | | 26.00 (2.97) | 24.00 (2.97) | 0.198 | | 0.595 | 0.805 | (0.571,1.14) | |
| | Missing/ No record | 55 | 18 | | | | | | |
| Absolute basophil | | 0.04 (0.01) | 0.04 (0.01) | 0.689 | | 0.935 | 1.06 | (0.753,1.42) | |
| | Missing/ No record | 35 | 14 | | | | | | |
| Absolute lymphocytes | | 1.75 (0.42) | 1.56 (0.50) | 0.143 | | 0.546 | 0.799 | (0.521,1.19) | |
| | Missing/ No record | 37 | 15 | | | | | | |
| Absolute neutrophils | | 3.66 (1.10) | 3.88 (1.10) | 0.074 | | 0.388 | 1.39 | (1.02,1.91) | |

| | | | | | | | | | |
|---|---|---|---|---|---|---|---|---|---|
| Absolute neutrophils | Missing/ No record | 35 | 14 | | | | | | |
| Segmented neutrophils | | 59.50 (8.30) | 61.80 (7.41) | 0.0552 | | 0.331 | 1.48 | (1.04,2.13) | |
| | Missing/ No record | 37 | 15 | | | | | | |
| Absolute eosinophils | | 0.17 (0.10) | 0.15 (0.11) | 0.369 | | 0.704 | 0.902 | (0.609,1.25) | |
| | Missing/ No record | 35 | 14 | | | | | | |
| Absolute monocytes | | 0.53 (0.13) | 0.53 (0.12) | 0.836 | | 1 | 1.13 | (0.833,1.48) | |
| | Missing/ No record | 35 | 14 | | | | | | |
| Platelet count | | 233.00 (54.86) | 222.50 (50.41) | 0.0328 | ** | 0.331 | 0.714 | (0.496,1.01) | 0.03 |
| | Missing/ No record | 36 | 14 | | | | | | |
| RBC | | 4.99 (0.40) | 4.91 (0.39) | 0.0521 | | 0.331 | 0.744 | (0.39,1.16) | |
| | Missing/ No record | 36 | 14 | | | | | | |
| RDW | | 12.80 (0.74) | 12.75 (0.59) | 0.88 | | 1 | 1.06 | (0.759,1.41) | |
| | Missing/ No record | 36 | 14 | | | | | | |
| MCH | | 30.10 (1.78) | 30.40 (1.11) | 0.273 | | 0.637 | 1.28 | (0.911,1.87) | |
| | Missing/ No record | 36 | 14 | | | | | | |
| MCHC | | 34.10 (0.89) | 34.00 (0.89) | 0.964 | | 1 | 1.02 | (0.74,1.41) | |
| | Missing/ No record | 36 | 14 | | | | | | |
| MCV | | 88.00 (4.45) | 89.00 (4.45) | 0.17 | | 0.569 | 1.39 | (0.974,2.04) | |
| | Missing/ No record | 36 | 14 | | | | | | |
| WBC | | 6.20 (1.78) | 6.40 (1.63) | 0.336 | | 0.704 | 1.39 | (1.05,2.14) | |
| | Missing/ No record | 36 | 14 | | | | | | |
| LMR | | 3.21 (1.07) | 2.93 (1.18) | 0.266 | | 0.637 | 0.793 | (0.44,1.29) | |
| | Missing/ No record | 37 | 15 | | | | | | |

**Supplementary Table S4c: L-CHIP-phenotype associations**

| Categorical Variable, N (%) | Characteristic | Negative (N=227) | L-CHIP positive (N=74) | p | | FDR | OR | 95% CI |
|---|---|---|---|---|---|---|---|---|
| Gender | F | 18 (0.0793) | 5 (0.0676) | 1 | | 1 | | |
| | M | 209 (0.921) | 69 (0.932) | | | | 1.19 | (0.455,3.71) |
| Smoking status | Never Smoked | 125 (0.551) | 36 (0.486) | 0.415 | | 0.909 | | |
| | Previous Smoker | 91 (0.401) | 36 (0.486) | | | | 1.37 | (0.804,2.35) |
| | Current Smoker | 11 (0.0485) | 2 (0.027) | | | | 0.631 | 0.0949,2.49) |
| Exposure | <= Low | 34 (0.15) | 12 (0.162) | 0.638 | | 0.992 | | |
| | Intermediate | 139 (0.612) | 46 (0.622) | | | | 0.938 | (0.458,2.02) |
| | >=High | 50 (0.22) | 12 (0.162) | | | | 0.68 | (0.271,1.7) |
| | Missing/ No record | 4 (0.0176) | 4 (0.0541) | | | | | |
| Race | Non-White | 18 (0.0793) | 8 (0.108) | 0.476 | | 0.909 | | |
| | White | 209 (0.921) | 66 (0.892) | | | | 0.711 | (0.304,1.8) |
| CVD | No | 207 (0.912) | 65 (0.878) | 0.374 | | 0.909 | | |
| | Yes | 20 (0.0881) | 9 (0.122) | | | | 1.43 | (0.595,3.22) |
| Stroke | No | 225 (0.991) | 73 (0.986) | 0.572 | | 0.925 | | |
| | Yes | 2 (0.00881) | 1 (0.0135) | | | | 1.54 | (0.071,16.3) |
| A | Het | 204 (0.899) | 63 (0.851) | 0.291 | | 0.909 | | |
| | Hom | 23 (0.101) | 11 (0.149) | | | | 1.55 | (0.692,3.29) |
| B | Het | 216 (0.952) | 72 (0.973) | 0.742 | | 1 | | |
| | Hom | 11 (0.0485) | 2 (0.027) | | | | 0.545 | 0.0831,2.09) |
| C | Het | 207 (0.912) | 70 (0.946) | 0.462 | | 0.909 | | |
| | Hom | 20 (0.0881) | 4 (0.0541) | | | | 0.591 | (0.168,1.63) |
| DRB1 | Het | 206 (0.907) | 71 (0.959) | 0.216 | | 0.908 | | |
| | Hom | 21 (0.0925) | 3 (0.0405) | | | | 0.414 | 0.0959,1.25) |
| DQA1 | Het | 202 (0.89) | 66 (0.892) | 1 | | 1 | | |
| | Hom | 25 (0.11) | 8 (0.108) | | | | 0.979 | (0.397,2.19) |
| DQB1 | Het | 206 (0.907) | 61 (0.824) | 0.0581 | | 0.848 | | |
| | Hom | 21 (0.0925) | 13 (0.176) | | | | 2.09 | (0.968,4.38) |
| DPA1 | Het | 78 (0.344) | 24 (0.324) | 0.888 | | 1 | | |
| | Hom | 149 (0.656) | 50 (0.676) | | | | 1.09 | (0.629,1.93) |
| DPB1 | Het | 179 (0.789) | 62 (0.838) | 0.406 | | 0.909 | | |
| | Hom | 48 (0.211) | 12 (0.162) | | | | 0.722 | (0.347,1.41) |
| DMA | Het | 63 (0.278) | 21 (0.284) | 1 | | 1 | | |
| | Hom | 164 (0.722) | 53 (0.716) | | | | 0.97 | (0.547,1.76) |
| DMB | Het | 119 (0.524) | 29 (0.392) | 0.0606 | | 0.848 | | |
| | Hom | 108 (0.476) | 45 (0.608) | | | | 1.71 | (1.01,2.94) |
| DOA | Het | 6 (0.0264) | 2 (0.027) | 1 | | 1 | | |
| | Hom | 221 (0.974) | 72 (0.973) | | | | 0.977 | (0.22,6.77) |
| DOB | Het | 95 (0.419) | 24 (0.324) | 0.172 | | 0.878 | | |
| | Hom | 132 (0.581) | 50 (0.676) | | | | 1.5 | (0.869,2.64) |
| DRA | Het | 98 (0.432) | 33 (0.446) | 0.893 | | 1 | | |
| | Hom | 129 (0.568) | 41 (0.554) | | | | 0.944 | (0.557,1.61) |

**Continuous Variable, Median (MAD)**

| | | | | | | | | |
|---|---|---|---|---|---|---|---|---|
| Age | | 59.00 (4.45) | 60.00 (4.45) | 0.098 | | 0.878 | 1.19 | (0.902,1.57) |
| | Missing/ No record | 0 | 0 | | | | | |
| BMI | | 30.11 (4.95) | 30.66 (4.63) | 0.768 | | 1 | 0.917 | (0.699,1.19) |
| | Missing/ No record | 0 | 0 | | | | | |
| Total cholesterol | | 190.50 (39.29) | 191.00 (37.81) | 0.994 | | 1 | 1.08 | (0.817,1.41) |
| | Missing/ No record | 35 | 8 | | | | | |
| Triglyceride | | 119.00 (60.05) | 117.00 (69.68) | 0.883 | | 1 | 0.913 | (0.675,1.2) |
| | Missing/ No record | 35 | 8 | | | | | |
| HDL | | 47.00 (11.86) | 49.50 (14.08) | 0.188 | | 0.878 | 1.28 | (0.977,1.69) |
| | Missing/ No record | 35 | 8 | | | | | |
| LDL | | 113.00 (32.62) | 107.00 (31.88) | 0.497 | | 0.909 | 0.906 | (0.681,1.2) |
| | Missing/ No record | 40 | 8 | | | | | |
| VLDL | | 24.00 (11.86) | 23.50 (14.08) | 0.855 | | 1 | 0.909 | (0.671,1.19) |
| | Missing/ No record | 35 | 8 | | | | | |

| | | | | | | | | |
|---|---|---|---|---|---|---|---|---|
| PCL | | 22.00 (7.41) | 25.75 (10.38) | 0.148 | | 0.878 | 1.06 | (0.82,1.36) |
| | Missing/ No record | 0 | 2 | | | | | |
| MoCA | | 26.00 (2.97) | 25.00 (2.97) | 0.0574 | | 0.848 | 0.77 | (0.574,1.03) |
| | Missing/ No record | 55 | 10 | | | | | |
| Absolute basophil | | 0.04 (0.01) | 0.04 (0.01) | 0.509 | | 0.909 | 1.15 | (0.878,1.49) |
| | Missing/ No record | 35 | 8 | | | | | |
| Absolute lymphocytes | | 1.75 (0.42) | 1.69 (0.53) | 0.981 | | 1 | 1.17 | (0.889,1.54) |
| | Missing/ No record | 37 | 9 | | | | | |
| Absolute neutrophils | | 3.66 (1.10) | 3.73 (0.98) | 0.46 | | 0.909 | 1.11 | (0.833,1.48) |
| | Missing/ No record | 35 | 8 | | | | | |
| Segmented neutrophils | | 59.50 (8.30) | 58.50 (8.90) | 0.802 | | 1 | 0.96 | (0.722,1.28) |
| | Missing/ No record | 37 | 9 | | | | | |
| Absolute eosinophils | | 0.17 (0.10) | 0.16 (0.10) | 0.374 | | 0.909 | 0.945 | (0.693,1.25) |
| | Missing/ No record | 35 | 9 | | | | | |
| Absolute monocytes | | 0.53 (0.13) | 0.54 (0.13) | 0.541 | | 0.909 | 1.12 | (0.854,1.45) |
| | Missing/ No record | 35 | 8 | | | | | |
| Platelet count | | 233.00 (54.86) | 225.50 (43.74) | 0.287 | | 0.909 | 0.893 | (0.672,1.18) |
| | Missing/ No record | 36 | 8 | | | | | |
| RBC | | 4.99 (0.40) | 4.89 (0.47) | 0.169 | | 0.878 | 0.9 | (0.53,1.2) |
| | Missing/ No record | 36 | 8 | | | | | |
| RDW | | 12.80 (0.74) | 12.85 (0.74) | 0.977 | | 1 | 0.995 | (0.744,1.29) |
| | Missing/ No record | 36 | 8 | | | | | |
| MCH | | 30.10 (1.78) | 30.25 (1.26) | 0.424 | | 0.909 | 1.14 | (0.861,1.53) |
| | Missing/ No record | 36 | 8 | | | | | |
| MCHC | | 34.10 (0.89) | 34.00 (0.59) | 0.47 | | 0.909 | 0.939 | (0.714,1.24) |
| | Missing/ No record | 36 | 8 | | | | | |
| MCV | | 88.00 (4.45) | 89.00 (2.97) | 0.181 | | 0.878 | 1.23 | (0.927,1.67) |
| | Missing/ No record | 36 | 8 | | | | | |
| WBC | | 6.20 (1.78) | 5.95 (1.78) | 0.539 | | 0.909 | 1.12 | (0.835,1.53) |
| | Missing/ No record | 36 | 8 | | | | | |
| LMR | | 3.21 (1.07) | 3.13 (1.19) | 0.747 | | 1 | 1.1 | (0.834,1.45) |
| | Missing/ No record | 37 | 9 | | | | | |

**Supplementary Table S4d: DNMT3A-phenotype associations**

| Categorical Variable, N (%) | Characteristic | Negative (N=227) | positive (N=22) | p | | FDR | OR | 95% CI | Multivariate GLM |
|---|---|---|---|---|---|---|---|---|---|
| Gender | F | 18 (0.0793) | 1 (0.0455) | 1 | | 1 | | | |
| | M | 209 (0.921) | 21 (0.955) | | | | 1.81 | (0.345,33.3) | |
| Smoking status | Never Smoked | 125 (0.551) | 8 (0.364) | 0.121 | | 0.568 | | | |
| | Previous Smoker | 91 (0.401) | 14 (0.636) | | | | 2.4 | (0.987,6.24) | |
| | Current Smoker | 11 (0.0485) | 0 (0) | | | | -- | -- | |
| Exposure | <= Low | 34 (0.15) | 7 (0.318) | 0.132 | | 0.568 | | | |
| | Intermediate | 139 (0.612) | 11 (0.5) | | | | 0.384 | (0.14,1.11) | |
| | >=High | 50 (0.22) | 3 (0.136) | | | | 0.291 | (0.0596,1.13) | |
| | Missing/ No record | 4 (0.0176) | 1 (0.0455) | | | | | | |
| Race | Non-White | 18 (0.0793) | 3 (0.136) | 0.411 | | 0.869 | | | |
| | White | 209 (0.921) | 19 (0.864) | | | | 0.545 | (0.165,2.47) | |
| CVD | No | 207 (0.912) | 21 (0.955) | 0.704 | | 0.948 | | | |
| | Yes | 20 (0.0881) | 1 (0.0455) | | | | 0.493 | (0.0268,2.56) | |
| Stroke | No | 225 (0.991) | 21 (0.955) | 0.243 | | 0.73 | | | |
| | Yes | 2 (0.00881) | 1 (0.0455) | | | | 5.36 | (0.243,58.2) | |
| A | Het | 204 (0.899) | 21 (0.955) | 0.705 | | 0.948 | | | |
| | Hom | 23 (0.101) | 1 (0.0455) | | | | 0.422 | (0.023,2.17) | |
| B | Het | 216 (0.952) | 22 (1) | 0.606 | | 0.948 | | | |
| | Hom | 11 (0.0485) | 0 (0) | | | | -- | -- | |
| C | Het | 207 (0.912) | 20 (0.909) | 1 | | 1 | | | |
| | Hom | 20 (0.0881) | 2 (0.0909) | | | | 1.03 | (0.158,3.91) | |
| DRB1 | Het | 206 (0.907) | 21 (0.955) | 0.703 | | 0.948 | | | |
| | Hom | 21 (0.0925) | 1 (0.0455) | | | | 0.467 | (0.0254,2.42) | |
| DQA1 | Het | 202 (0.89) | 19 (0.864) | 0.722 | | 0.948 | | | |
| | Hom | 25 (0.11) | 3 (0.136) | | | | 1.28 | (0.286,4.09) | |
| DQB1 | Het | 206 (0.907) | 20 (0.909) | 1 | | 1 | | | |
| | Hom | 21 (0.0925) | 2 (0.0909) | | | | 0.981 | (0.15,3.69) | |
| DPA1 | Het | 78 (0.344) | 8 (0.364) | 0.819 | | 0.992 | | | |
| | Hom | 149 (0.656) | 14 (0.636) | | | | 0.916 | (0.376,2.38) | |
| DPB1 | Het | 179 (0.789) | 17 (0.773) | 0.791 | | 0.992 | | | |
| | Hom | 48 (0.211) | 5 (0.227) | | | | 1.1 | (0.347,2.94) | |
| DMA | Het | 63 (0.278) | 2 (0.0909) | 0.0737 | | 0.568 | | | |
| | Hom | 164 (0.722) | 20 (0.909) | | | | 3.84 | (1.08,24.5) | |
| DMB | Het | 119 (0.524) | 10 (0.455) | 0.656 | | 0.948 | | | |
| | Hom | 108 (0.476) | 12 (0.545) | | | | 1.32 | (0.549,3.25) | |
| DOA | Het | 6 (0.0264) | 0 (0) | 1 | | 1 | | | |
| | Hom | 221 (0.974) | 22 (1) | | | | -- | -- | |
| DOB | Het | 95 (0.419) | 7 (0.318) | 0.497 | | 0.869 | | | |
| | Hom | 132 (0.581) | 15 (0.682) | | | | 1.54 | (0.625,4.17) | |
| DRA | Het | 98 (0.432) | 10 (0.455) | 0.826 | | 0.992 | | | |
| | Hom | 129 (0.568) | 12 (0.545) | | | | 0.912 | (0.378,2.24) | |

**Continuous Variable, Median (MAD)**

| | | | | | | | | | |
|---|---|---|---|---|---|---|---|---|---|
| Age | | 59.00 (4.45) | 66.50 (9.64) | 3.82E-05 | ** | 0.00161 | 2.3 | (1.58,3.44) | 0.0132 |
| | Missing/ No record | 0 | 0 | | | | | | |
| BMI | | 30.11 (4.95) | 27.30 (2.77) | 0.0173 | ** | 0.364 | 0.539 | (0.307,0.888) | 0.994 |
| | Missing/ No record | 0 | 0 | | | | | | |
| Total cholesterol | | 190.50 (39.29) | 177.00 (53.37) | 0.344 | | 0.849 | 0.649 | (0.335,1.19) | |
| | Missing/ No record | 35 | 7 | | | | | | |
| Triglyceride | | 119.00 (60.05) | 94.00 (48.93) | 0.149 | | 0.568 | 0.582 | (0.235,1.11) | |
| | Missing/ No record | 35 | 7 | | | | | | |
| HDL | | 47.00 (11.86) | 52.00 (13.34) | 0.128 | | 0.568 | 1.46 | (0.829,2.48) | |
| | Missing/ No record | 35 | 7 | | | | | | |
| LDL | | 113.00 (32.62) | 98.00 (31.13) | 0.183 | | 0.608 | 0.671 | (0.375,1.15) | |
| | Missing/ No record | 40 | 7 | | | | | | |
| VLDL | | 24.00 (11.86) | 19.00 (10.38) | 0.144 | | 0.568 | 0.58 | (0.234,1.11) | |
| | Missing/ No record | 35 | 7 | | | | | | |
| PCL | | 22.00 (7.41) | 21.50 (6.67) | 0.277 | | 0.775 | 0.618 | (0.32,1.04) | |
| | Missing/ No record | 0 | 0 | | | | | | |
| MoCA | | 26.00 (2.97) | 24.00 (2.22) | 0.188 | | 0.608 | 0.747 | (0.428,1.35) | |
| | Missing/ No record | 55 | 10 | | | | | | |
| Absolute basophil | | 0.04 (0.01) | 0.04 (0.01) | 0.593 | | 0.948 | 0.739 | (0.357,1.36) | |
| | Missing/ No record | 35 | 7 | | | | | | |
| Absolute lymphocytes | | 1.75 (0.42) | 1.46 (0.37) | 0.0469 | ** | 0.568 | 0.469 | (0.198,0.989) | 0.14 |
| | Missing/ No record | 37 | 8 | | | | | | |
| Absolute neutrophils | | 3.66 (1.10) | 3.52 (1.07) | 0.911 | | 1 | 1.09 | (0.629,1.79) | |
| | Missing/ No record | 35 | 7 | | | | | | |
| Segmented neutrophils | | 59.50 (8.30) | 61.10 (6.15) | 0.424 | | 0.869 | 1.29 | (0.732,2.32) | |
| | Missing/ No record | 37 | 8 | | | | | | |
| Absolute eosinophils | | 0.17 (0.10) | 0.16 (0.09) | 0.343 | | 0.849 | 0.604 | (0.249,1.18) | |
| | Missing/ No record | 35 | 7 | | | | | | |
| Absolute monocytes | | 0.53 (0.13) | 0.54 (0.16) | 0.656 | | 0.948 | 1.29 | (0.925,1.76) | |
| | Missing/ No record | 35 | 7 | | | | | | |
| Platelet count | | 233.00 (54.86) | 225.00 (35.58) | 0.0929 | | 0.568 | 0.589 | (0.324,1.02) | |

| | | | | | | | | |
|---|---|---|---|---|---|---|---|---|
| Platelet count | Missing/ No record | 36 | 7 | | | | | |
| RBC | | 4.99 (0.40) | 4.92 (0.33) | 0.426 | | 0.869 | 0.92 | (0.376,1.32) |
| | Missing/ No record | 36 | 7 | | | | | |
| RDW | | 12.80 (0.74) | 12.50 (0.59) | 0.455 | | 0.869 | 0.751 | (0.343,1.33) |
| | Missing/ No record | 36 | 7 | | | | | |
| MCH | | 30.10 (1.78) | 29.80 (1.93) | 0.91 | | 1 | 1.14 | (0.698,1.99) |
| | Missing/ No record | 36 | 7 | | | | | |
| MCHC | | 34.10 (0.89) | 34.00 (1.04) | 0.467 | | 0.869 | 0.835 | (0.526,1.37) |
| | Missing/ No record | 36 | 7 | | | | | |
| MCV | | 88.00 (4.45) | 88.00 (4.45) | 0.488 | | 0.869 | 1.36 | (0.793,2.46) |
| | Missing/ No record | 36 | 7 | | | | | |
| WBC | | 6.20 (1.78) | 6.20 (1.78) | 0.975 | | 1 | 1.34 | (0.996,1.99) |
| | Missing/ No record | 36 | 7 | | | | | |
| LMR | | 3.21 (1.07) | 2.76 (0.84) | 0.0908 | | 0.568 | 0.355 | (0.101,1.02) |
| | Missing/ No record | 37 | 8 | | | | | |

**Supplementary Table S4e: TET2-phenotype associations**

| Categorical Variable, N (%) | Characteristic | Negative (N=227) | TET2 positive (N=15) | p | | FDR | OR | 95% CI | Multivariate GLM |
|---|---|---|---|---|---|---|---|---|---|
| Gender | F | 18 (0.0793) | 1 (0.0667) | 1 | | 1 | | | |
| | M | 209 (0.921) | 14 (0.933) | | | | 1.21 | (0.222,22.5) | |
| Smoking status | Never Smoked | 125 (0.551) | 6 (0.4) | 0.295 | | 0.687 | | | |
| | Previous Smoker | 91 (0.401) | 9 (0.6) | | | | 2.06 | (0.718,6.34) | |
| | Current Smoker | 11 (0.0485) | 0 (0) | | | | -- | -- | |
| Exposure | <= Low | 34 (0.15) | 2 (0.133) | 0.361 | | 0.687 | | | |
| | Intermediate | 139 (0.612) | 12 (0.8) | | | | 1.47 | (0.377,9.71) | |
| | >=High | 50 (0.22) | 1 (0.0667) | | | | 0.34 | (0.0154,3.68) | |
| | Missing/ No record | 4 (0.0176) | 0 (0) | | | | | | |
| Race | Non-White | 18 (0.0793) | 3 (0.2) | 0.13 | | 0.634 | | | |
| | White | 209 (0.921) | 12 (0.8) | | | | 0.344 | (0.0981,1.61) | |
| CVD | No | 207 (0.912) | 13 (0.867) | 0.633 | | 0.792 | | | |
| | Yes | 20 (0.0881) | 2 (0.133) | | | | 1.59 | (0.238,6.32) | |
| Stroke | No | 225 (0.991) | 15 (1) | 1 | | 1 | | | |
| | Yes | 2 (0.00881) | 0 (0) | | | | -- | -- | |
| A | Het | 204 (0.899) | 14 (0.933) | 1 | | 1 | | | |
| | Hom | 23 (0.101) | 1 (0.0667) | | | | 0.634 | (0.0342,3.38) | |
| B | Het | 216 (0.952) | 14 (0.933) | 0.545 | | 0.789 | | | |
| | Hom | 11 (0.0485) | 1 (0.0667) | | | | 1.4 | (0.0741,8.04) | |
| C | Het | 207 (0.912) | 14 (0.933) | 1 | | 1 | | | |
| | Hom | 20 (0.0881) | 1 (0.0667) | | | | 0.739 | (0.0398,3.98) | |
| DRB1 | Het | 206 (0.907) | 15 (1) | 0.375 | | 0.687 | | | |
| | Hom | 21 (0.0925) | 0 (0) | | | | -- | -- | |
| DQA1 | Het | 202 (0.89) | 12 (0.8) | 0.393 | | 0.687 | | | |
| | Hom | 25 (0.11) | 3 (0.2) | | | | 2.02 | (0.439,6.9) | |
| DQB1 | Het | 206 (0.907) | 13 (0.867) | 0.641 | | 0.792 | | | |
| | Hom | 21 (0.0925) | 2 (0.133) | | | | 1.51 | (0.226,5.97) | |
| DPA1 | Het | 78 (0.344) | 8 (0.533) | 0.166 | | 0.634 | | | |
| | Hom | 149 (0.656) | 7 (0.467) | | | | 0.458 | (0.155,1.32) | |
| DPB1 | Het | 179 (0.789) | 14 (0.933) | 0.317 | | 0.687 | | | |
| | Hom | 48 (0.211) | 1 (0.0667) | | | | 0.266 | (0.0145,1.38) | |
| DMA | Het | 63 (0.278) | 7 (0.467) | 0.142 | | 0.634 | | | |
| | Hom | 164 (0.722) | 8 (0.533) | | | | 0.439 | (0.151,1.3) | |
| DMB | Het | 119 (0.524) | 4 (0.267) | 0.0642 | | 0.539 | | | |
| | Hom | 108 (0.476) | 11 (0.733) | | | | 3.03 | (1,11.2) | |
| DOA | Het | 6 (0.0264) | 0 (0) | 1 | | 1 | | | |
| | Hom | 221 (0.974) | 15 (1) | | | | -- | -- | |
| DOB | Het | 95 (0.419) | 10 (0.667) | 0.104 | | 0.634 | | | |
| | Hom | 132 (0.581) | 5 (0.333) | | | | 0.36 | (0.109,1.05) | |
| DRA | Het | 98 (0.432) | 8 (0.533) | 0.592 | | 0.792 | | | |
| | Hom | 129 (0.568) | 7 (0.467) | | | | 0.665 | (0.226,1.91) | |

**Continuous Variable, Median (MAD)**

| | | | | | | | | | |
|---|---|---|---|---|---|---|---|---|---|
| Age | | 59.00 (4.45) | 64.00 (8.90) | 0.00701 | ** | 0.295 | 1.87 | (1.2,2.94) | 0.00335 |
| | Missing/ No record | 0 | 0 | | | | | | |
| BMI | | 30.11 (4.95) | 30.41 (4.60) | 0.98 | | 1 | 0.982 | (0.57,1.59) | |
| | Missing/ No record | 0 | 0 | | | | | | |
| Total cholesterol | | 190.50 (39.29) | 186.00 (38.55) | 0.611 | | 0.792 | 0.8 | (0.372,1.59) | |
| | Missing/ No record | 35 | 4 | | | | | | |
| Triglyceride | | 119.00 (60.05) | 153.00 (72.65) | 0.375 | | 0.687 | 1.07 | (0.565,1.72) | |
| | Missing/ No record | 35 | 4 | | | | | | |
| HDL | | 47.00 (11.86) | 45.00 (11.86) | 0.601 | | 0.792 | 1.01 | (0.473,1.91) | |
| | Missing/ No record | 35 | 4 | | | | | | |
| LDL | | 113.00 (32.62) | 94.00 (29.65) | 0.373 | | 0.687 | 0.757 | (0.387,1.41) | |
| | Missing/ No record | 40 | 4 | | | | | | |
| VLDL | | 24.00 (11.86) | 31.00 (14.83) | 0.385 | | 0.687 | 1.07 | (0.564,1.72) | |
| | Missing/ No record | 35 | 4 | | | | | | |
| PCL | | 22.00 (7.41) | 28.00 (14.83) | 0.477 | | 0.765 | 1.15 | (0.697,1.75) | |
| | Missing/ No record | 0 | 0 | | | | | | |
| MoCA | | 26.00 (2.97) | 24.00 (2.97) | 0.401 | | 0.687 | 0.779 | (0.474,1.33) | |
| | Missing/ No record | 55 | 1 | | | | | | |
| Absolute basophil | | 0.04 (0.01) | 0.04 (0.01) | 0.403 | | 0.687 | 0.706 | (0.295,1.43) | |
| | Missing/ No record | 35 | 4 | | | | | | |
| Absolute lymphocytes | | 1.75 (0.42) | 1.47 (0.24) | 0.0453 | ** | 0.478 | 0.474 | (0.182,1.07) | 0.586 |
| | Missing/ No record | 37 | 4 | | | | | | |
| Absolute neutrophils | | 3.66 (1.10) | 4.12 (0.76) | 0.382 | | 0.687 | 1.26 | (0.678,2.2) | |
| | Missing/ No record | 35 | 4 | | | | | | |
| Segmented neutrophils | | 59.50 (8.30) | 66.10 (4.60) | 0.0456 | ** | 0.478 | 1.86 | (0.968,3.82) | 0.393 |
| | Missing/ No record | 37 | 4 | | | | | | |

| | | | | | | | | | |
|---|---|---|---|---|---|---|---|---|---|
| Absolute eosinophils | | 0.17 (0.10) | 0.09 (0.13) | 0.386 | | 0.687 | 0.743 | (0.298,1.41) | |
| | Missing/ No record | 35 | 4 | | | | | | |
| Absolute monocytes | | 0.53 (0.13) | 0.50 (0.10) | 0.264 | | 0.687 | 0.55 | (0.164,1.32) | |
| | Missing/ No record | 35 | 4 | | | | | | |
| Platelet count | | 233.00 (54.86) | 210.00 (38.55) | 0.16 | | 0.634 | 0.669 | (0.337,1.25) | |
| | Missing/ No record | 36 | 4 | | | | | | |
| RBC | | 4.99 (0.40) | 4.62 (0.49) | 0.0406 | ** | 0.478 | 0.715 | (0.27,1.31) | 0.735 |
| | Missing/ No record | 36 | 4 | | | | | | |
| RDW | | 12.80 (0.74) | 13.10 (0.89) | 0.136 | | 0.634 | 1.22 | (0.666,1.9) | |
| | Missing/ No record | 36 | 4 | | | | | | |
| MCH | | 30.10 (1.78) | 29.90 (1.33) | 0.909 | | 1 | 1.01 | (0.6,1.9) | |
| | Missing/ No record | 36 | 4 | | | | | | |
| MCHC | | 34.10 (0.89) | 33.80 (0.59) | 0.409 | | 0.687 | 0.84 | (0.493,1.5) | |
| | Missing/ No record | 36 | 4 | | | | | | |
| MCV | | 88.00 (4.45) | 88.00 (4.45) | 0.951 | | 1 | 1.1 | (0.626,2.14) | |
| | Missing/ No record | 36 | 4 | | | | | | |
| WBC | | 6.20 (1.78) | 6.60 (1.78) | 0.517 | | 0.776 | 1.65 | (1.07,2.68) | |
| | Missing/ No record | 36 | 4 | | | | | | |
| LMR | | 3.21 (1.07) | 3.14 (0.90) | 0.492 | | 0.765 | 0.658 | (0.192,1.59) | |
| | Missing/ No record | 37 | 4 | | | | | | |

**Supplementary Table S4f: PPM1D-phenotype associations**

| Categorical Variable, N (%) | Characteristic | Negative (N=227) | PPM1D positive (N=11) | p | | FDR | OR | 95% CI | Multivariate GLM |
|---|---|---|---|---|---|---|---|---|---|
| Gender | F | 18 (0.0793) | 1 (0.0909) | 1 | | 1 | | | |
| | M | 209 (0.921) | 10 (0.909) | | | | 0.861 | (0.152,16.2) | |
| Smoking status | Never Smoked | 125 (0.551) | 4 (0.364) | 0.281 | | 0.656 | | | |
| | Previous Smoker | 91 (0.401) | 6 (0.545) | | | | 2.06 | (0.572,8.26) | |
| | Current Smoker | 11 (0.0485) | 1 (0.0909) | | | | 2.84 | (0.139,21.4) | |
| Exposure | <= Low | 34 (0.15) | 3 (0.273) | 0.192 | | 0.656 | | | |
| | Intermediate | 139 (0.612) | 4 (0.364) | | | | 0.326 | (0.0688,1.72) | |
| | >=High | 50 (0.22) | 4 (0.364) | | | | 0.907 | (0.188,4.84) | |
| | Missing/ No record | 4 (0.0176) | 0 (0) | | | | | | |
| Race | Non-White | 18 (0.0793) | 1 (0.0909) | 1 | | 1 | | | |
| | White | 209 (0.921) | 10 (0.909) | | | | 0.861 | (0.152,16.2) | |
| CVD | No | 207 (0.912) | 9 (0.818) | 0.27 | | 0.656 | | | |
| | Yes | 20 (0.0881) | 2 (0.182) | | | | 2.3 | (0.336,9.72) | |
| Stroke | No | 225 (0.991) | 11 (1) | 1 | | 1 | | | |
| | Yes | 2 (0.00881) | 0 (0) | | | | -- | -- | |
| A | Het | 204 (0.899) | 8 (0.727) | 0.106 | | 0.538 | | | |
| | Hom | 23 (0.101) | 3 (0.273) | | | | 3.33 | (0.693,12.4) | |
| B | Het | 216 (0.952) | 10 (0.909) | 0.441 | | 0.842 | | | |
| | Hom | 11 (0.0485) | 1 (0.0909) | | | | 1.96 | (0.102,11.7) | |
| C | Het | 207 (0.912) | 10 (0.909) | 1 | | 1 | | | |
| | Hom | 20 (0.0881) | 1 (0.0909) | | | | 1.03 | (0.055,5.83) | |
| DRB1 | Het | 206 (0.907) | 10 (0.909) | 1 | | 1 | | | |
| | Hom | 21 (0.0925) | 1 (0.0909) | | | | 0.981 | (0.0522,5.51) | |
| DQA1 | Het | 202 (0.89) | 7 (0.636) | 0.0322 | ** | 0.538 | | | 0.146 |
| | Hom | 25 (0.11) | 4 (0.364) | | | | 4.62 | (1.14,16.4) | |
| DQB1 | Het | 206 (0.907) | 8 (0.727) | 0.0867 | | 0.538 | | | |
| | Hom | 21 (0.0925) | 3 (0.273) | | | | 3.68 | (0.763,13.8) | |
| DPA1 | Het | 78 (0.344) | 3 (0.273) | 0.754 | | 1 | | | |
| | Hom | 149 (0.656) | 8 (0.727) | | | | 1.4 | (0.391,6.51) | |
| DPB1 | Het | 179 (0.789) | 7 (0.636) | 0.262 | | 0.656 | | | |
| | Hom | 48 (0.211) | 4 (0.364) | | | | 2.13 | (0.54,7.36) | |
| DMA | Het | 63 (0.278) | 3 (0.273) | 1 | | 1 | | | |
| | Hom | 164 (0.722) | 8 (0.727) | | | | 1.02 | (0.286,4.79) | |
| DMB | Het | 119 (0.524) | 6 (0.545) | 1 | | 1 | | | |
| | Hom | 108 (0.476) | 5 (0.455) | | | | 0.918 | (0.258,3.13) | |
| DOA | Het | 6 (0.0264) | 0 (0) | 1 | | 1 | | | |
| | Hom | 221 (0.974) | 11 (1) | | | | -- | -- | |
| DOB | Het | 95 (0.419) | 4 (0.364) | 1 | | 1 | | | |
| | Hom | 132 (0.581) | 7 (0.636) | | | | 1.26 | (0.37,4.92) | |
| DRA | Het | 98 (0.432) | 6 (0.545) | 0.54 | | 0.938 | | | |
| | Hom | 129 (0.568) | 5 (0.455) | | | | 0.633 | (0.178,2.16) | |

**Continuous Variable, Median (MAD)**

| | | | | | | | | |
|---|---|---|---|---|---|---|---|---|
| Age | | 59.00 (4.45) | 60.00 (4.45) | 0.369 | | 0.813 | 1.39 | (0.78,2.25) |
| | Missing/ No record | 0 | 0 | | | | | |
| BMI | | 30.11 (4.95) | 29.41 (3.94) | 0.267 | | 0.656 | 0.564 | (0.264,1.09) |
| | Missing/ No record | 0 | 0 | | | | | |
| Total cholesterol | | 190.50 (39.29) | 195.00 (28.17) | 1 | | 1 | 0.909 | (0.399,1.89) |
| | Missing/ No record | 35 | 2 | | | | | |
| Triglyceride | | 119.00 (60.05) | 127.00 (31.13) | 0.407 | | 0.813 | 0.984 | (0.448,1.7) |
| | Missing/ No record | 35 | 2 | | | | | |
| HDL | | 47.00 (11.86) | 50.00 (13.34) | 0.595 | | 0.938 | 1.29 | (0.602,2.51) |
| | Missing/ No record | 35 | 2 | | | | | |
| LDL | | 113.00 (32.62) | 105.00 (41.51) | 0.603 | | 0.938 | 0.821 | (0.397,1.62) |
| | Missing/ No record | 40 | 2 | | | | | |
| VLDL | | 24.00 (11.86) | 25.00 (7.41) | 0.403 | | 0.813 | 0.982 | (0.446,1.69) |
| | Missing/ No record | 35 | 2 | | | | | |
| PCL | | 22.00 (7.41) | 22.00 (4.26) | 0.943 | | 1 | 0.695 | (0.284,1.32) |
| | Missing/ No record | 0 | 0 | | | | | |
| MoCA | | 26.00 (2.97) | 24.00 (2.97) | 0.14 | | 0.538 | 0.597 | (0.301,1.23) |
| | Missing/ No record | 55 | 4 | | | | | |
| Absolute basophil | | 0.04 (0.01) | 0.04 (0.01) | 0.917 | | 1 | 0.984 | (0.424,1.87) |
| | Missing/ No record | 35 | 2 | | | | | |
| Absolute lymphocytes | | 1.75 (0.42) | 1.64 (0.49) | 0.576 | | 0.938 | 1.06 | (0.471,2.12) |
| | Missing/ No record | 37 | 2 | | | | | |
| Absolute neutrophils | | 3.66 (1.10) | 3.98 (0.95) | 0.0988 | | 0.538 | 1.61 | (0.855,2.92) |
| | Missing/ No record | 35 | 2 | | | | | |
| Segmented neutrophils | | 59.50 (8.30) | 61.80 (5.63) | 0.24 | | 0.656 | 1.63 | (0.811,3.45) |
| | Missing/ No record | 37 | 2 | | | | | |
| Absolute eosinophils | | 0.17 (0.10) | 0.20 (0.22) | 0.66 | | 0.99 | 1.34 | (0.766,2.06) |

| | | | | | | | | | |
|---|---|---|---|---|---|---|---|---|---|
| Absolute eosinophils | Missing/ No record | 35 | 2 | | | | | | |
| Absolute monocytes | | 0.53 (0.13) | 0.54 (0.15) | 0.792 | | 1 | 1.01 | (0.359,1.59) | |
| | Missing/ No record | 35 | 2 | | | | | | |
| Platelet count | | 233.00 (54.86) | 183.00 (50.41) | 0.0497 | ** | 0.538 | 0.474 | (0.208,0.982) | 0.117 |
| | Missing/ No record | 36 | 2 | | | | | | |
| RBC | | 4.99 (0.40) | 5.02 (0.42) | 0.538 | | 0.938 | 0.901 | (0.302,1.37) | |
| | Missing/ No record | 36 | 2 | | | | | | |
| RDW | | 12.80 (0.74) | 12.50 (0.30) | 0.141 | | 0.538 | 0.475 | (0.135,1.2) | |
| | Missing/ No record | 36 | 2 | | | | | | |
| MCH | | 30.10 (1.78) | 31.00 (1.78) | 0.049 | ** | 0.538 | 2.22 | (1.07,4.91) | 0.172 |
| | Missing/ No record | 36 | 2 | | | | | | |
| MCHC | | 34.10 (0.89) | 34.70 (0.44) | 0.0739 | | 0.538 | 1.76 | (0.886,3.82) | |
| | Missing/ No record | 36 | 2 | | | | | | |
| MCV | | 88.00 (4.45) | 90.00 (5.93) | 0.135 | | 0.538 | 2.06 | (0.995,4.47) | |
| | Missing/ No record | 36 | 2 | | | | | | |
| WBC | | 6.20 (1.78) | 6.40 (0.59) | 0.113 | | 0.538 | 1.24 | (0.362,2.24) | |
| | Missing/ No record | 36 | 2 | | | | | | |
| LMR | | 3.21 (1.07) | 2.57 (0.85) | 0.223 | | 0.656 | 1.04 | (0.354,2.04) | |
| | Missing/ No record | 37 | 2 | | | | | | |

**Supplementary Table S4g: EEF1A1-phenotype associations**

| Categorical Variable, N (%) | Characteristic | Negative (N=227) | EEF1A1 positive (N=18) | p | | FDR | OR | 95% CI |
|---|---|---|---|---|---|---|---|---|
| Gender | F | 18 (0.0793) | 2 (0.111) | 0.648 | | 1 | | |
| | M | 209 (0.921) | 16 (0.889) | | | | 0.689 | (0.176,4.57) |
| Smoking status | Never Smoked | 125 (0.551) | 11 (0.611) | 1 | | 1 | | |
| | Previous Smoker | 91 (0.401) | 7 (0.389) | | | | 0.874 | (0.311,2.31) |
| | Current Smoker | 11 (0.0485) | 0 (0) | | | | -- | -- |
| Exposure | <= Low | 34 (0.15) | 3 (0.167) | 0.0545 | | 0.846 | | |
| | Intermediate | 139 (0.612) | 14 (0.778) | | | | 1.14 | (0.348,5.15) |
| | >=High | 50 (0.22) | 0 (0) | | | | -- | -- |
| | Missing/ No record | 4 (0.0176) | 1 (0.0556) | | | | | |
| Race | Non-White | 18 (0.0793) | 3 (0.167) | 0.191 | | 0.846 | | |
| | White | 209 (0.921) | 15 (0.833) | | | | 0.431 | (0.127,1.98) |
| CVD | No | 207 (0.912) | 18 (1) | 0.375 | | 0.868 | | |
| | Yes | 20 (0.0881) | 0 (0) | | | | -- | -- |
| Stroke | No | 225 (0.991) | 18 (1) | 1 | | 1 | | |
| | Yes | 2 (0.00881) | 0 (0) | | | | -- | -- |
| A | Het | 204 (0.899) | 16 (0.889) | 1 | | 1 | | |
| | Hom | 23 (0.101) | 2 (0.111) | | | | 1.11 | (0.169,4.24) |
| B | Het | 216 (0.952) | 16 (0.889) | 0.246 | | 0.846 | | |
| | Hom | 11 (0.0485) | 2 (0.111) | | | | 2.45 | (0.36,10.2) |
| C | Het | 207 (0.912) | 17 (0.944) | 1 | | 1 | | |
| | Hom | 20 (0.0881) | 1 (0.0556) | | | | 0.609 | (0.0329,3.22) |
| DRB1 | Het | 206 (0.907) | 16 (0.889) | 0.68 | | 1 | | |
| | Hom | 21 (0.0925) | 2 (0.111) | | | | 1.23 | (0.186,4.72) |
| DQA1 | Het | 202 (0.89) | 16 (0.889) | 1 | | 1 | | |
| | Hom | 25 (0.11) | 2 (0.111) | | | | 1.01 | (0.154,3.84) |
| DQB1 | Het | 206 (0.907) | 15 (0.833) | 0.398 | | 0.868 | | |
| | Hom | 21 (0.0925) | 3 (0.167) | | | | 1.96 | (0.43,6.57) |
| DPA1 | Het | 78 (0.344) | 12 (0.667) | 0.00969 | ** | 0.407 | | |
| | Hom | 149 (0.656) | 6 (0.333) | | | | 0.262 | (0.0882,0.701) |
| DPB1 | Het | 179 (0.789) | 17 (0.944) | 0.135 | | 0.846 | | |
| | Hom | 48 (0.211) | 1 (0.0556) | | | | 0.219 | (0.012,1.11) |
| DMA | Het | 63 (0.278) | 5 (0.278) | 1 | | 1 | | |
| | Hom | 164 (0.722) | 13 (0.722) | | | | 0.999 | (0.36,3.22) |
| DMB | Het | 119 (0.524) | 6 (0.333) | 0.145 | | 0.846 | | |
| | Hom | 108 (0.476) | 12 (0.667) | | | | 2.2 | (0.826,6.52) |
| DOA | Het | 6 (0.0264) | 1 (0.0556) | 0.418 | | 0.868 | | |
| | Hom | 221 (0.974) | 17 (0.944) | | | | 0.462 | (0.0728,8.98) |
| DOB | Het | 95 (0.419) | 8 (0.444) | 1 | | 1 | | |
| | Hom | 132 (0.581) | 10 (0.556) | | | | 0.9 | (0.342,2.44) |
| DRA | Het | 98 (0.432) | 8 (0.444) | 1 | | 1 | | |
| | Hom | 129 (0.568) | 10 (0.556) | | | | 0.95 | (0.361,2.57) |

**Continuous Variable, Median (MAD)**

| | | | | | | | | |
|---|---|---|---|---|---|---|---|---|
| Age | | 59.00 (4.45) | 59.00 (3.71) | 0.887 | | 1 | 0.945 | (0.517,1.55) |
| | Missing/ No record | 0 | 0 | | | | | |
| BMI | | 30.11 (4.95) | 30.80 (4.04) | 0.626 | | 1 | 1.02 | (0.625,1.6) |
| | Missing/ No record | 0 | 0 | | | | | |
| Total cholesterol | | 190.50 (39.29) | 182.50 (41.51) | 0.8 | | 1 | 0.928 | (0.52,1.59) |
| | Missing/ No record | 35 | 0 | | | | | |
| Triglyceride | | 119.00 (60.05) | 110.00 (49.67) | 0.322 | | 0.846 | 0.671 | (0.318,1.16) |
| | Missing/ No record | 35 | 0 | | | | | |
| HDL | | 47.00 (11.86) | 52.50 (12.60) | 0.128 | | 0.846 | 1.59 | (1.06,2.42) |
| | Missing/ No record | 35 | 0 | | | | | |
| LDL | | 113.00 (32.62) | 99.50 (33.36) | 0.434 | | 0.868 | 0.813 | (0.483,1.33) |
| | Missing/ No record | 40 | 0 | | | | | |
| VLDL | | 24.00 (11.86) | 22.00 (9.64) | 0.308 | | 0.846 | 0.669 | (0.316,1.16) |
| | Missing/ No record | 35 | 0 | | | | | |
| PCL | | 22.00 (7.41) | 26.78 (10.70) | 0.265 | | 0.846 | 0.985 | (0.583,1.51) |
| | Missing/ No record | 0 | 0 | | | | | |
| MoCA | | 26.00 (2.97) | 25.00 (4.45) | 0.152 | | 0.846 | 0.715 | (0.443,1.17) |

| | | | | | | | | |
|---|---|---|---|---|---|---|---|---|
| MoCA | Missing/ No record | 55 | 1 | | | | | |
| Absolute basophil | | 0.04 (0.01) | 0.04 (0.01) | 0.92 | | 1 | 0.927 | (0.501,1.54) |
| | Missing/ No record | 35 | 0 | | | | | |
| Absolute lymphocytes | | 1.75 (0.42) | 1.74 (0.43) | 0.801 | | 1 | 1.12 | (0.627,1.89) |
| | Missing/ No record | 37 | 0 | | | | | |
| Absolute neutrophils | | 3.66 (1.10) | 3.25 (1.17) | 0.31 | | 0.846 | 0.784 | (0.447,1.3) |
| | Missing/ No record | 35 | 0 | | | | | |
| Segmented neutrophils | | 59.50 (8.30) | 56.30 (6.52) | 0.205 | | 0.846 | 0.75 | (0.458,1.23) |
| | Missing/ No record | 37 | 0 | | | | | |
| Absolute eosinophils | | 0.17 (0.10) | 0.15 (0.10) | 0.262 | | 0.846 | 0.773 | (0.383,1.3) |
| | Missing/ No record | 35 | 0 | | | | | |
| Absolute monocytes | | 0.53 (0.13) | 0.55 (0.10) | 0.536 | | 0.938 | 1.07 | (0.582,1.56) |
| | Missing/ No record | 35 | 0 | | | | | |
| Platelet count | | 233.00 (54.86) | 221.50 (52.63) | 0.308 | | 0.846 | 0.76 | (0.451,1.24) |
| | Missing/ No record | 36 | 0 | | | | | |
| RBC | | 4.99 (0.40) | 4.86 (0.47) | 0.369 | | 0.868 | 0.957 | (0.433,1.32) |
| | Missing/ No record | 36 | 0 | | | | | |
| RDW | | 12.80 (0.74) | 12.90 (0.96) | 0.922 | | 1 | 0.85 | (0.446,1.38) |
| | Missing/ No record | 36 | 0 | | | | | |
| MCH | | 30.10 (1.78) | 30.35 (1.19) | 0.752 | | 1 | 1.1 | (0.708,1.84) |
| | Missing/ No record | 36 | 0 | | | | | |
| MCHC | | 34.10 (0.89) | 33.85 (0.67) | 0.513 | | 0.938 | 0.947 | (0.606,1.53) |
| | Missing/ No record | 36 | 0 | | | | | |
| MCV | | 88.00 (4.45) | 89.50 (2.97) | 0.517 | | 0.938 | 1.17 | (0.731,1.97) |
| | Missing/ No record | 36 | 0 | | | | | |
| WBC | | 6.20 (1.78) | 5.40 (2.45) | 0.067 | | 0.846 | 0.179 | (0.0326,0.928) |
| | Missing/ No record | 36 | 0 | | | | | |
| LMR | | 3.21 (1.07) | 2.99 (1.11) | 0.6 | | 1 | 0.901 | (0.39,1.65) |
| | Missing/ No record | 37 | 0 | | | | | |

**Supplementary Table S4h: DDX11-phenotype associations**

| Categorical Variable, N (%) | Characteristic | Negative (N=227) | positive (N=13) | p | | FDR | OR | 95% CI | Multivariate GLM |
|---|---|---|---|---|---|---|---|---|---|
| Gender | F | 18 (0.0793) | 3 (0.231) | 0.0932 | | 0.486 | | | |
| | M | 209 (0.921) | 10 (0.769) | | | | 0.287 | (0.0791,1.36) | |
| Smoking status | Never Smoked | 125 (0.551) | 4 (0.308) | 0.17 | | 0.596 | | | |
| | Previous Smoker | 91 (0.401) | 8 (0.615) | | | | 2.75 | (0.839,10.5) | |
| | Current Smoker | 11 (0.0485) | 1 (0.0769) | | | | 2.84 | (0.139,21.4) | |
| Exposure | <= Low | 34 (0.15) | 4 (0.308) | 0.199 | | 0.644 | | | |
| | Intermediate | 139 (0.612) | 7 (0.538) | | | | 0.428 | (0.122,1.71) | |
| | >=High | 50 (0.22) | 1 (0.0769) | | | | 0.17 | (0.00849,1.21) | |
| | Missing/ No record | 4 (0.0176) | 1 (0.0769) | | | | | | |
| Race | Non-White | 18 (0.0793) | 2 (0.154) | 0.296 | | 0.79 | | | |
| | White | 209 (0.921) | 11 (0.846) | | | | 0.474 | (0.115,3.21) | |
| CVD | No | 207 (0.912) | 11 (0.846) | 0.339 | | 0.79 | | | |
| | Yes | 20 (0.0881) | 2 (0.154) | | | | 1.88 | (0.279,7.66) | |
| Stroke | No | 225 (0.991) | 13 (1) | 1 | | 1 | | | |
| | Yes | 2 (0.00881) | 0 (0) | | | | -- | -- | |
| A | Het | 204 (0.899) | 11 (0.846) | 0.632 | | 0.948 | | | |
| | Hom | 23 (0.101) | 2 (0.154) | | | | 1.61 | (0.24,6.5) | |
| B | Het | 216 (0.952) | 12 (0.923) | 0.496 | | 0.922 | | | |
| | Hom | 11 (0.0485) | 1 (0.0769) | | | | 1.64 | (0.086,9.54) | |
| C | Het | 207 (0.912) | 11 (0.846) | 0.339 | | 0.79 | | | |
| | Hom | 20 (0.0881) | 2 (0.154) | | | | 1.88 | (0.279,7.66) | |
| DRB1 | Het | 206 (0.907) | 13 (1) | 0.612 | | 0.948 | | | |
| | Hom | 21 (0.0925) | 0 (0) | | | | -- | -- | |
| DQA1 | Het | 202 (0.89) | 12 (0.923) | 1 | | 1 | | | |
| | Hom | 25 (0.11) | 1 (0.0769) | | | | 0.673 | (0.0362,3.64) | |
| DQB1 | Het | 206 (0.907) | 11 (0.846) | 0.36 | | 0.795 | | | |
| | Hom | 21 (0.0925) | 2 (0.154) | | | | 1.78 | (0.265,7.24) | |
| DPA1 | Het | 78 (0.344) | 3 (0.231) | 0.551 | | 0.948 | | | |
| | Hom | 149 (0.656) | 10 (0.769) | | | | 1.74 | (0.516,7.95) | |
| DPB1 | Het | 179 (0.789) | 10 (0.769) | 1 | | 1 | | | |
| | Hom | 48 (0.211) | 3 (0.231) | | | | 1.12 | (0.244,3.83) | |
| DMA | Het | 63 (0.278) | 3 (0.231) | 1 | | 1 | | | |
| | Hom | 164 (0.722) | 10 (0.769) | | | | 1.28 | (0.377,5.85) | |
| DMB | Het | 119 (0.524) | 5 (0.385) | 0.399 | | 0.837 | | | |
| | Hom | 108 (0.476) | 8 (0.615) | | | | 1.76 | (0.571,5.99) | |
| DOA | Het | 6 (0.0264) | 1 (0.0769) | 0.326 | | 0.79 | | | |
| | Hom | 221 (0.974) | 12 (0.923) | | | | 0.326 | (0.0498,6.41) | |
| DOB | Het | 95 (0.419) | 3 (0.231) | 0.249 | | 0.748 | | | |
| | Hom | 132 (0.581) | 10 (0.769) | | | | 2.4 | (0.712,10.9) | |
| DRA | Het | 98 (0.432) | 5 (0.385) | 0.783 | | 1 | | | |
| | Hom | 129 (0.568) | 8 (0.615) | | | | 1.22 | (0.393,4.13) | |

**Continuous Variable, Median (MAD)**

| | | | | | | | | | |
|---|---|---|---|---|---|---|---|---|---|
| Age | | 59.00 (4.45) | 59.00 (2.97) | 0.662 | | 0.958 | 0.989 | (0.492,1.72) | |
| | Missing/ No record | 0 | 0 | | | | | | |
| BMI | | 30.11 (4.95) | 31.17 (4.69) | 0.632 | | 0.948 | 1.16 | (0.677,1.9) | |
| | Missing/ No record | 0 | 0 | | | | | | |
| Total cholesterol | | 190.50 (39.29) | 197.00 (65.23) | 0.927 | | 1 | 0.91 | (0.469,1.67) | |
| | Missing/ No record | 35 | 0 | | | | | | |
| Triglyceride | | 119.00 (60.05) | 118.00 (75.61) | 0.883 | | 1 | 0.925 | (0.471,1.51) | |
| | Missing/ No record | 35 | 0 | | | | | | |
| HDL | | 47.00 (11.86) | 52.00 (19.27) | 0.116 | | 0.486 | 1.67 | (0.944,2.88) | |
| | Missing/ No record | 35 | 0 | | | | | | |
| LDL | | 113.00 (32.62) | 115.00 (26.69) | 0.448 | | 0.897 | 0.746 | (0.409,1.32) | |
| | Missing/ No record | 40 | 0 | | | | | | |
| VLDL | | 24.00 (11.86) | 24.00 (16.31) | 0.908 | | 1 | 0.919 | (0.466,1.51) | |
| | Missing/ No record | 35 | 0 | | | | | | |
| PCL | | 22.00 (7.41) | 28.00 (11.86) | 0.0468 | ** | 0.33 | 1.32 | (0.811,2.03) | 0.779 |
| | Missing/ No record | 0 | 0 | | | | | | |
| MoCA | | 26.00 (2.97) | 22.00 (2.97) | 0.00241 | ** | 0.101 | 0.413 | (0.227,0.722) | 0.00657 |
| | Missing/ No record | 55 | 2 | | | | | | |
| Absolute basophil | | 0.04 (0.01) | 0.04 (0.01) | 0.984 | | 1 | 0.937 | (0.459,1.66) | |
| | Missing/ No record | 35 | 0 | | | | | | |
| Absolute lymphocytes | | 1.75 (0.42) | 2.07 (0.71) | 0.0114 | ** | 0.239 | 2.15 | (1.23,3.8) | 0.0807 |
| | Missing/ No record | 37 | 0 | | | | | | |

| | | | | | | | | | |
|---|---|---|---|---|---|---|---|---|---|
| Absolute neutrophils | | 3.66 (1.10) | 3.85 (1.41) | 0.881 | | 1 | 1.06 | (0.585,1.82) | |
| | Missing/ No record | 35 | 0 | | | | | | |
| Segmented neutrophils | | 59.50 (8.30) | 51.90 (6.97) | 0.0487 | ** | 0.33 | 0.622 | (0.353,1.08) | 0.53 |
| | Missing/ No record | 37 | 0 | | | | | | |
| Absolute eosinophils | | 0.17 (0.10) | 0.16 (0.15) | 0.87 | | 1 | 1.12 | (0.604,1.75) | |
| | Missing/ No record | 35 | 1 | | | | | | |
| Absolute monocytes | | 0.53 (0.13) | 0.55 (0.12) | 0.505 | | 0.922 | 1.01 | (0.436,1.54) | |
| | Missing/ No record | 35 | 0 | | | | | | |
| Platelet count | | 233.00 (54.86) | 229.00 (57.82) | 0.823 | | 1 | 0.913 | (0.505,1.6) | |
| | Missing/ No record | 36 | 0 | | | | | | |
| RBC | | 4.99 (0.40) | 4.71 (0.36) | 0.107 | | 0.486 | 0.861 | (0.33,1.32) | |
| | Missing/ No record | 36 | 0 | | | | | | |
| RDW | | 12.80 (0.74) | 12.80 (0.44) | 0.611 | | 0.948 | 0.687 | (0.282,1.31) | |
| | Missing/ No record | 36 | 0 | | | | | | |
| MCH | | 30.10 (1.78) | 30.80 (0.89) | 0.0551 | | 0.33 | 1.91 | (1.03,3.73) | |
| | Missing/ No record | 36 | 0 | | | | | | |
| MCHC | | 34.10 (0.89) | 34.00 (0.59) | 0.752 | | 1 | 0.971 | (0.586,1.68) | |
| | Missing/ No record | 36 | 0 | | | | | | |
| MCV | | 88.00 (4.45) | 91.00 (5.93) | 0.0192 | ** | 0.269 | 2.37 | (1.25,4.75) | 0.0758 |
| | Missing/ No record | 36 | 0 | | | | | | |
| WBC | | 6.20 (1.78) | 6.80 (1.78) | 0.148 | | 0.564 | 1.15 | (0.339,2.07) | |
| | Missing/ No record | 36 | 0 | | | | | | |
| LMR | | 3.21 (1.07) | 4.05 (0.80) | 0.0286 | ** | 0.3 | 1.54 | (0.833,2.7) | 0.877 |
| | Missing/ No record | 37 | 0 | | | | | | |
